# Unraveling the shared genetics of common epilepsies and general cognitive ability

**DOI:** 10.1101/2024.03.25.24304773

**Authors:** Naz Karadag, Espen Hagen, Alexey A. Shadrin, Dennis van der Meer, Kevin S. O’Connell, Zillur Rahman, Gleda Kutrolli, Nadine Parker, Shahram Bahrami, Vera Fominykh, Kjell Heuser, Erik Taubøll, Torill Ueland, Nils Eiel Steen, Srdjan Djurovic, Anders M. Dale, Oleksandr Frei, Ole A. Andreassen, Olav B. Smeland

**Author notes:** **Corresponding Author:** Olav B. Smeland, Centre for Precision Psychiatry, Division of Mental Health and Addiction University of Oslo and Oslo University Hospital, Kirkeveien 166, 0424 Oslo, Norway. Shared first-author (Naz Karadag, Espen Hagen).

## Abstract

**Objective:** Cognitive impairment is prevalent among individuals with epilepsy, and it is possible that genetic factors can underlie this relationship. Here, we investigated the potential shared genetic basis of common epilepsies and general cognitive ability (COG).

**Methods:** We applied linkage disequilibrium score (LDSC) regression, MiXeR and conjunctional false discovery rate (conjFDR) to analyze different aspects of genetic overlap between COG and epilepsies. We used the largest available genome-wide association study data on COG (*n* = 269,867) and common epilepsies (*n* = 27,559 cases, 42,436 controls), including the broad phenotypes ‘all epilepsy’, focal epilepsies and genetic generalized epilepsies (GGE), and as well as specific subtypes. We functionally annotated the identified loci using a variety of biological resources and validated the results in independent samples.

**Results:** Using MiXeR, COG (11.2k variants) was estimated to be almost four times more polygenic than ‘all epilepsy’, GGE, juvenile myoclonic epilepsy (JME), and childhood absence epilepsy (CAE) (2.5k – 2.9k variants). The other epilepsy phenotypes were insufficiently powered for analysis. We show extensive genetic overlap between COG and epilepsies with significant negative genetic correlations (-0.23 to -0.04). COG was estimated to share 2.9k variants with both GGE and ‘all epilepsy’, and 2.3k variants with both JME and CAE. Using conjFDR, we identified 66 distinct loci shared between COG and epilepsies, including novel associations for GGE (27), ‘all epilepsy’ (5), JME (5) and CAE (5). The implicated genes were significantly expressed in multiple brain regions. The results were validated in independent samples (COG: *p* = 1.0 x 10^-14^; ‘all epilepsy’: *p* = 5.6 x 10^-3^).

**Significance:** Our study demonstrates a substantial genetic basis shared between epilepsies and COG and identifies novel overlapping genomic loci. Enhancing our understanding of the relationship between epilepsies and COG may lead to the development of novel comorbidity-targeted epilepsy treatments.

## Introduction

Epilepsies are diverse brain disorders marked by unprovoked recurrent seizures.^1^ Epilepsies contribute significantly to the global disease burden, affecting over 60 million people worldwide of all ages.^2^ The underlying pathogenesis of epilepsies remains poorly understood and many patients continue to suffer from uncontrolled seizures.^3,4^ Epilepsies are divided by seizure onset into two broad groups; focal epilepsies and generalized epilepsies, the latter being predominantly composed of genetic generalized epilepsies (GGE). While focal epilepsies and GGE differ in their clinical presentations, both are associated with cognitive impairments, which substantially affect the quality of life of individuals with epilepsies.^5,6^ Cognitive impairments of individuals with epilepsies encompass difficulties with memory, attention, language and executive functioning, which in many cases predate seizure onset, emphasizing that cognitive impairment is not merely a consequence of seizures.^5^ However, not all individuals with epilepsy have poor cognitive functioning, a wide range of cognitive performance among individuals with epilepsy suggests a complex relationship between epilepsies and cognition.^5,7^ Poor cognitive ability is also associated with several clinical and etiological characteristics of epilepsy, including the origin, frequency and duration of seizures, age of seizure onset, duration of epilepsy, cerebral pathology, and effect of anti-seizure medication.^8^ Further, studies have suggested that cognitive performance, particularly in the domains of intelligence and memory, has prognostic value for seizure outcomes in patients with epilepsies.^9–11^ While surgical intervention may improve cognitive functioning in a selected group of focal epilepsy patients, anti-seizure medication is rather associated with a worsening of cognitive function by suppressing neuronal excitability or enhancing inhibitory neurotransmission.^3,12,13^ Increasing evidence favors a bidirectional relationship between epilepsies and cognition, as part of a pathogenesis altering neural networks thereby leading to both cognitive impairments and the propensity of seizures.^14,15^ A better understanding of the relationship between epilepsies and cognition may help the development of novel comorbidity-targeted epilepsy treatments.

Both cognitive abilities and epilepsies are heritable;^16–19^ suggesting that shared genetic underpinnings could contribute to the observed cognitive deficits in epilepsies. Previous studies have reported that the heritability attributable to common genetic variants is 19% for general cognitive ability (COG) and 9% and 32% for focal epilepsy and GGE, respectively.^16,19^ Recent genome-wide association studies (GWAS) have identified over 200 genomic risk loci associated with COG and over 60 loci associated with epilepsies.^16–20^ Furthermore, significant negative genetic correlations between COG and epilepsies (‘all epilepsy’, focal epilepsies and GGE) have been reported (*r*_g_ = -0.20, -0.23, -0.14, respectively), indicating shared genetic effects underlying these phenotypes.^16,17,21–23^ Moreover, recent modeling work using bivariate MiXeR^24^ has estimated that a considerable fraction of the genetic architecture of GGE overlaps with COG.^17^ However, to what extent other epilepsy subtypes share genetic variants with COG remains poorly understood and the specific loci jointly involved in these phenotypes are mostly unknown. Identification of these loci could inform the specific molecular genetic mechanisms shared between COG and epilepsies.

To gain more insights into the shared genetic basis of COG and epilepsies including several epilepsy subtypes beyond the broad categories, we analyzed the largest available GWAS samples on COG and three broad epilepsy types ‘all epilepsy’, focal epilepsies and GGE, and their seven subtypes, utilizing the tools linkage disequilibrium score regression (LDSC)^25^, MiXeR^24^ and conjunctional false discovery rate (conjFDR).^26,27^ While LDSC^25^ estimates pairwise global genetic correlations, MiXeR^24^ estimates the number of variants underlying each phenotype as well as the number of variants shared between them, irrespective of the genetic correlations. In addition, conjFDR improves the discovery of overlapping genomic loci^26,27^ and has facilitated the identification of an extensive list of genetic variants shared between various complex human phenotypes regardless of their allelic effect directions,^28,29^ including epilepsies and COG with other traits.^18,30–32^

## Methods

### Sample Description

#### Discovery samples

We obtained GWAS data (Table 1) in the form of summary statistics (*p*-values and effect sizes). All participants were of European ancestry to ensure compatible patterns of linkage disequilibrium (LD). The datasets were controlled for systemic sample overlap to avoid potential bias. The summary statistics on COG^19^ (*n* = 269,867) were based on a GWAS meta-analysis from 14 cohorts, with UK Biobank being the largest cohort. The cohorts either reported Spearman’s *g* underlying several dimensions of cognitive functioning or a primary measure of fluid intelligence that correlates highly with *g*.^19^

**TABLE 1.**
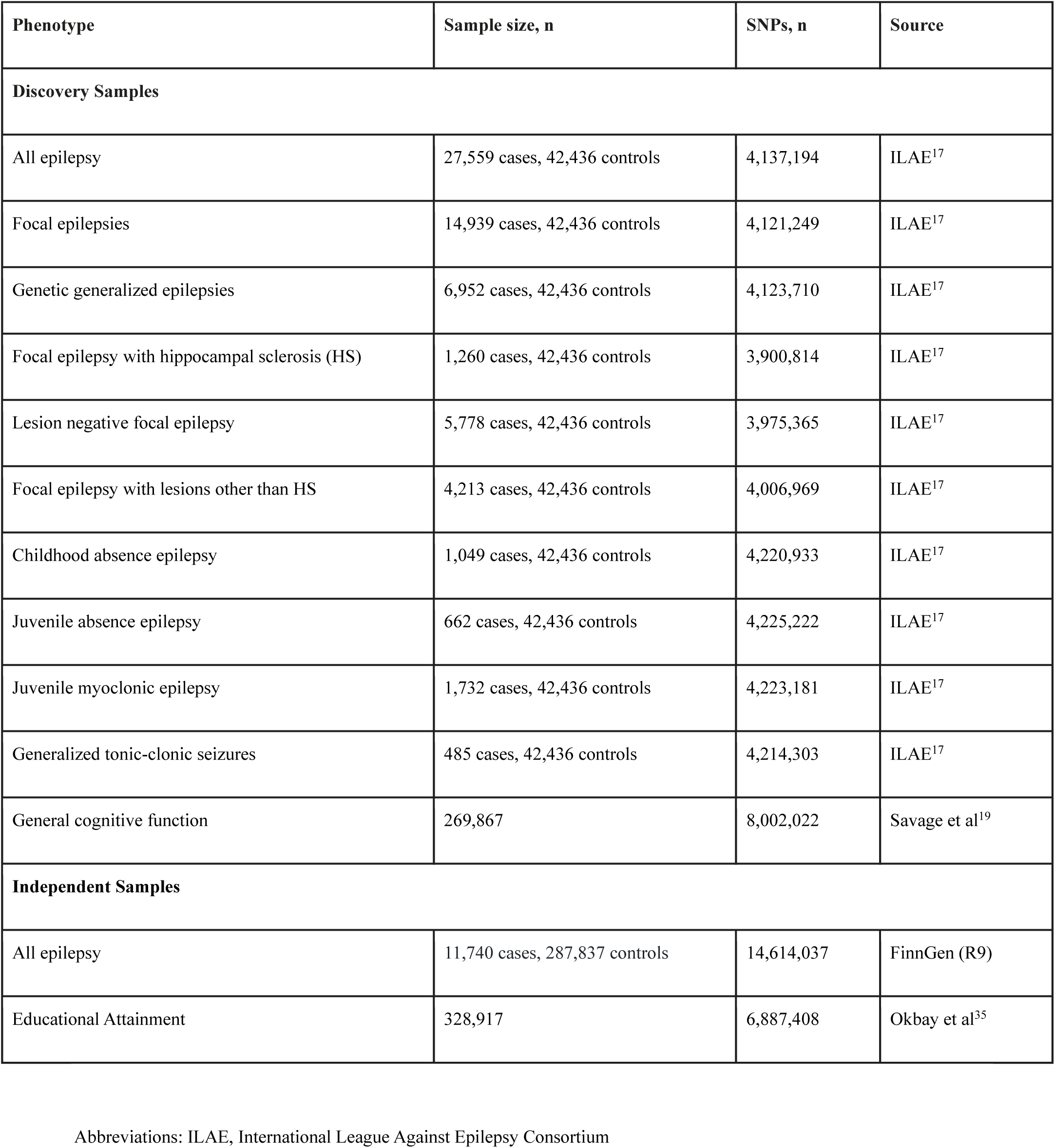
Summary Data from all GWAS used in the present study.

| Phenotype | Sample size, n | SNPs, n | Source |
| --- | --- | --- | --- |
| **Discovery Samples** |  |  |  |
| All epilepsy | 27,559 cases, 42,436 controls | 4,137,194 | ILAE<sup>17</sup> |
| Focal epilepsies | 14,939 cases, 42,436 controls | 4,121,249 | ILAE<sup>17</sup> |
| Genetic generalized epilepsies | 6,952 cases, 42,436 controls | 4,123,710 | ILAE<sup>17</sup> |
| Focal epilepsy with hippocampal sclerosis (HS) | 1,260 cases, 42,436 controls | 3,900,814 | ILAE<sup>17</sup> |
| Lesion negative focal epilepsy | 5,778 cases, 42,436 controls | 3,975,365 | ILAE<sup>17</sup> |
| Focal epilepsy with lesions other than HS | 4,213 cases, 42,436 controls | 4,006,969 | ILAE<sup>17</sup> |
| Childhood absence epilepsy | 1,049 cases, 42,436 controls | 4,220,933 | ILAE<sup>17</sup> |
| Juvenile absence epilepsy | 662 cases, 42,436 controls | 4,225,222 | ILAE<sup>17</sup> |
| Juvenile myoclonic epilepsy | 1,732 cases, 42,436 controls | 4,223,181 | ILAE<sup>17</sup> |
| Generalized tonic-clonic seizures | 485 cases, 42,436 controls | 4,214,303 | ILAE<sup>17</sup> |
| General cognitive function | 269,867 | 8,002,022 | Savage et al<sup>19</sup> |
| **Independent Samples** |  |  |  |
| All epilepsy | 11,740 cases, 287,837 controls | 14,614,037 | FinnGen (R9) |
| Educational Attainment | 328,917 | 6,887,408 | Okbay et al<sup>35</sup> |

Abbreviations: ILAE, International League Against Epilepsy Consortium

The summary statistics for epilepsies^17^ (*n* = 27,559 cases, 42,436 controls) were acquired from the International League Against Epilepsy (ILAE) Consortium and included data on the broad phenotypes ‘all epilepsy’, focal epilepsies and GGE as well as the focal subtypes ‘lesion negative focal epilepsy’, ‘focal epilepsy with hippocampal sclerosis’, ‘focal epilepsy with lesions other than hippocampal sclerosis’ and the generalized subtypes childhood absence epilepsy (CAE), juvenile absence epilepsy, juvenile myoclonic epilepsy (JME) and generalized tonic-clonic seizures.

All GWAS summary statistics underwent quality control and were formatted with the cleansumstats pipeline v1.6.0.^33^

#### Independent samples

To assess the reliability of our results, we also acquired GWAS summary statistics from independent samples. The independent epilepsies data was collected from FinnGen (freeze 9; https://r9.finngen.fi r9, phenotypic code G6_EPILEPSY) and consists of all epilepsies as a combined phenotype (*n* = 11,740 cases, 287,837 controls).^34^ We used an independent sample^35^ on educational attainment as a proxy for COG, as they are highly genetically correlated (*r*_g_ = 0.73).^19^ All independent samples were based on participants of European ancestry in accordance with the discovery samples.

#### Ethics statement

All GWAS investigated in the present study were approved by the relevant ethics committees, and informed consent was obtained from all participants. The Regional Committee for Medical and Health Research Ethics for the South-East Norway found that no additional institutional review board approval was needed.

### Data Analysis

#### Filtering of summary statistics

To avoid inflation due to LD, SNPs in the extended major histocompatibility complex (MHC) region, chromosome 8p23.1 and the microtubule-associated protein tau (MAPT) region (genome build GRCh37/hg19 locations chr6:25119106-33854733; chr8:7200000-12500000; chr17:40000000-47000000, respectively) were excluded in all analyses before fitting the statistical models.^27,36^

#### Linkage disequilibrium score regression analysis

LDSC^25^ was applied to estimate pairwise genetic correlations (*r*_g_). LDSC estimates genetic correlations at the genome-wide level and distinguishes between the contributions from polygenic effects and confounding factors. Multiple testing correction was carried out by the Benjamini-Hochberg method (*q* < 0.05).

#### Gaussian causal mixture models

We utilized univariate and bivariate Gaussian causal mixture models to GWAS data using MiXeR (v1.3).^24^ With maximum likelihood estimation, univariate MiXeR estimates the distribution of SNPs with non-null additive genetic effects beyond LD quantifying the number of causal variants explaining 90% of the SNP-heritability (polygenicity), and the variance of the effect sizes of the SNPs with non-null genetic effects (discoverability). Then, the SNP-heritability is computed on the observed scale based on the polygenicity and discoverability estimates. Further, bivariate MiXeR estimates the number of overlapping and phenotype-specific causal variants between two phenotypes.

All point estimates and standard deviations were computed by performing 20 iterations with 2 million randomly selected SNPs which were then further randomly pruned at LD (*r*^2^) threshold of 0.8, (resulting in ∼600K SNPs per iteration). Akaike information criterion (AIC) and log-likelihood plots were evaluated for the model fit. See the original publication for more information on MiXeR.^24^

#### Conjunctional false discovery rate analysis

We applied the conjFDR analysis implemented in the pleioFDR software tool to increase genetic discovery of the shared loci between epilepsies and COG.^26,27^ The conjFDR analysis is an extension to the conditional false discovery rate (condFDR), which readjusts the test statistics in a primary phenotype (e.g., GGE) by conditioning on SNP associations with a secondary phenotype (e.g., COG). The conjFDR performs two condFDR analyses by first conditioning the primary phenotype on the secondary phenotype and then conditioning the secondary phenotype on the primary phenotype and selects the conjFDR value as the maximum of the two condFDR values. The conjFDR threshold of 0.05 was used in line with previous studies.^26,27^ Using a linkage disequilibrium (LD) threshold *r*^2^ = 0.01, SNPs were randomly pruned over 500 iterations to avoid bias in the analysis.^27^

The cross-trait enrichment is visualized by conditional quantile-quantile (Q-Q) plots, which plot p-value distributions for a primary phenotype for all SNPs, and for SNP strata defined by their association with a secondary phenotype. A strong cross-trait enrichment is observed by a leftward deflection from the null hypothesis (diagonal) with a decrease in p-values in the Q-Q plots. See the original publications for more detailed information on condFDR/conjFDR methods.^26,27^

#### Sign concordance test

To validate the conjFDR findings *en masse*, we applied the sign concordance test in independent samples. We assessed the overall pattern of concordance in allelic effect directions of the lead SNPs of all identified loci between the discovery and the independent samples. For loci shared with more than one epilepsy phenotype, we used the most strongly associated lead SNP only. The point-estimates of the beta coefficients were compared to determine how many lead SNPs in the shared loci had concordant allelic directions in the discovery and the independent samples. Under the null hypothesis assumption, given that there is no genetic association with the trait of interest, the likelihood of randomly detecting sign concordance is 50%. We assessed if the observed sign concordance rates were significantly higher than expected (more than 50%) by a two-tailed exact binomial test.

### Functional Analyses

#### Genomic loci definition

Genomic loci were defined as independent if independent significant SNPs had *r*^2^<0.60 and conjFDR<0.05 in accordance with the Functional Mapping and Annotation of GWAS (FUMA) platform.^37^ Lead SNPs were subsequently identified as independent significant SNPs with *r*^2^ < 0.1 in approximate LD. All SNPs with conjFDR < 0.10 and in LD (*r*^2^ > 0.60) with an independent significant SNP were chosen as the candidate SNPs. For merged loci within >250kb of each other, the SNP with the most significant conjFDR value was chosen as the lead SNP of the merged locus. Candidate SNPs in LD (*r*^2^ ≥ 0.6) with one of the independent SNPs in the locus defined the locus borders. All LD *r*^2^ values were obtained from the 1000 Genomes Project European-ancestry haplotype reference panel.^38^

The concordance effects of the shared loci were evaluated by studying their z-scores and odds ratios. Novel loci were identified as loci at a minimum distance of 500kb from the reported loci from the original GWAS or not listed by the GWAS Catalog^39^ or OpenTargets Genetics^40^ or other GWAS analyses on epilepsies or COG.

#### Functional annotation

Combined annotation dependent depletion scores (CADD), regulomeDB scores and chromatin state scores were used to functionally annotate the candidate SNPs, in line with FUMA.^37^ CADD score predicts deleterious SNP effects on proteins, regulomeDB score predicts the probability of a SNP to have a regulatory function and chromatin state scores predict transcriptional effects. The Variant-to-GENE (V2G) pipeline from the open-source OpenTargets Genetics was utilized to map the lead SNPs to likely causal genes.^40^ V2G employs information on the distance of SNPs to genes, chromatin interactions and molecular phenotype quantitative trait loci investigations (QTL) and uses this information in machine learning algorithms for gene mapping for each lead SNP. Gene expression and gene set analyses for the GO, KEGG and canonical pathways gene sets were carried out as hypergeometric tests using FUMA and Genotype-Tissue Expression data (GTEx) on the input of the identified genes from V2G.^41,42^ The mapped genes were investigated using the National Center for Biotechnology Information database.^43^ Using Brain RNA-Seq,^44^ we then leveraged the single-cell RNA sequencing (RNA-Seq) data to assess whether the mapped genes were significantly expressed in brain cells.

## Results

### Linkage disequilibrium score regression analysis

We estimated the SNP-heritability of each phenotype and the genetic correlations between all pairs of phenotypes using LDSC^25^ (Supplementary Table 1). We estimated SNP-heritabilities of 0.19 for COG (*h*^2^ SE= 6.80 x 10^-3^), 0.12 for ‘all epilepsy’ (*h*^2^ SE= 0.01), 0.07 for focal epilepsies (*h*^2^ SE= 0.01) and 0.60 for GGE (*h*^2^ SE= 0.05). We also observed that focal epilepsy subtypes (*h*^2^ range = 0.03 – 0.25) had lower estimated SNP-heritabilities compared to GGE subtypes (*h*^2^ range = 0.60 – 0.93), while juvenile absence epilepsy and generalized tonic-clonic seizures were insufficiently powered for the analysis. Further, we found that five epilepsy phenotypes (‘all epilepsy’, focal epilepsies, ‘lesion negative focal epilepsies’, GGE and JME) were significantly negatively correlated with COG, in line with previous findings.^16,21,22^ The significant genetic correlations between COG and ‘lesion negative focal epilepsies’ (*r*_g_ = -0.23, SE = 0.07, *p* = 3.00 x 10^-4^) and between COG and JME (*r*_g_ = -0.12, SE = 0.04, *p* = 3.30 x 10^-3^) are novel reports to our knowledge. The genetic correlations between COG and ‘focal epilepsies with hippocampal sclerosis’, ‘focal epilepsies with lesions other than hippocampal sclerosis’ and CAE did not reach significance.

### MiXeR analysis

Univariate MiXeR^24^ analyses were sufficiently powered for ‘all epilepsy’, GGE, CAE, JME and COG only, as indicated by positive AIC scores (Supplementary Table 2). COG was estimated to be almost four times more polygenic than the four epilepsy phenotypes, which had similar polygenicities. Specifically, COG was estimated to be influenced by 11k (SD = 0.3k) variants, ‘all epilepsy’ by 2.9k (SD = 0.4k) variants, GGE by 2.9k (SD = 0.2k) variants, JME by 2.5k (SD = 0.3k) variants and CAE by 2.5k (SD = 0.4k) variants.

We then conducted bivariate MiXeR and estimated the overlapping genomic proportion between COG and the four sufficiently powered epilepsy phenotypes and observed almost complete overlap with COG (Supplementary Table 3). 2.9k (SD = 0.4k) variants were estimated to be shared between COG and ‘all epilepsy’, 2.9k (SD = 0.2k) between COG and GGE, 2.3k (SD = 0.4k) between COG and CAE, and 2.3k (SD = 0.3k) between COG and JME. The fraction of concordant variants within the overlapping variants was 0.42 for COG and GGE (SD = 6.9 x 10^-3^) and for COG and CAE (SD = 1.6 x 10^-2^), 0.41 for COG and JME (SD = 1.1 x 10^-2^) and 0.37 for COG and ‘all epilepsy’ (SD = 8.4 x 10^-3^); indicating mixed allelic effect directions among the overlapping variants, but an overabundance (concordance rate below 0.50) of epilepsy risk variants linked to worse cognitive performance.

### Conjunctional false discovery rate analysis

In line with the LDSC and MiXeR results, we observed substantial cross-trait enrichment between COG and the same four epilepsy phenotypes (GGE, JME, CAE and ‘all epilepsy’), as visualized by the conditional quantile-quantile (Q-Q) plots (Supplementary Figure 2). No cross-trait enrichment was observed for any of the focal epilepsies, generalized tonic-clonic seizures or juvenile absence epilepsy.

Next, we leveraged the cross-trait enrichment using conjFDR^26,27^ analysis to boost the discovery of overlapping genomic loci (Supplementary tables 4-7). We identified shared loci between COG and ‘all epilepsy’ (11), GGE (55), CAE (9) and JME (11). 15 of the loci were linked to at least two epilepsy phenotypes where three loci were overlapping in three or more analyses. Specifically, one locus at chromosome 2 near the gene *CTD-2026C7.1* was shared between ‘all epilepsy’, GGE, CAE and COG; another locus at chromosome 5 near *RP11- 492A10.1* was shared between the generalized epilepsies GGE, CAE, JME and COG; and lastly, one locus at chromosome 10 near *C10orf76* was shared between all four epilepsy phenotypes and COG. In summary, we found 27 novel loci for GGE, five novel loci for ‘all epilepsy’, five novel loci for JME and five novel loci for CAE. We did not find any novel loci for COG as the shared loci have been identified previously by other studies.^19,20^ Notably, we also found a locus within the MAPT region. The MAPT region involves a complex LD structure, covering many genes, with genetic variants linked to neurodegenerative disorders, learning disability and brain morphology.^45^ Therefore, our finding only indicates that MAPT is involved in epilepsies and COG, instead of a specific gene or locus.

We determined the allelic effect directions of the lead SNPs for each locus. 21 out of 55 lead SNPs for GGE, one out of 11 lead SNPs for ‘all epilepsy’, five out of nine lead SNPs for CAE and four out of 11 lead SNPs shared with JME had concordant allelic effect directions.

### Sign concordance test

For the distinct epilepsy loci identified at conjFDR < 0.05, 42 out of 63 (67%) lead SNPs were sign concordant in the independent epilepsy sample^34^ (binomial test *p* = 5.57 x 10^−3^; Supplementary Table 8). For COG, 62 out of 66 (94%) lead SNPs were sign concordant in the independent proxy sample^35^ (*p* = 1.04 x 10^−14^).

### Functional annotation

Functional annotation of the candidate SNPs indicated that most of them are positioned in intronic and intergenic regions (Supplementary Tables 9-12). There was a total of 23 nonsynonymous exonic variants located across 14 loci (Supplementary Table 13). Among the implicated genes were *LONRF2*, *STAB1*, *GNL3*, *ITIH1*, *CTD-2117L12.1*, *ELL2*, *RMI1*, *IER5L*, *FBXO3*, *SERPING1*, *DDN*, *TNRC6A*, *PER1*, *UPK1A*, *ZNFX1* and *BRWD1*. Some of these genes have previously been identified by other studies on epilepsies, in particular *STAB1*, *GNL3*, *ITIH1*, *ELL2*, *RMI1, DDN* and *UPK1A*.^16–18,46^ Across analyses, 63 candidate SNPs had a CADD-score higher than 12.37 which indicates deleteriousness.^47^ We applied OpenTargets to map genes to each lead SNP based on their V2G scores (Supplementary Tables 14-17). We further performed hypergeometric tests as implemented in FUMA for gene expression and gene set analyses of the identified genes from OpenTargets. The mapped genes were significantly enriched for expression in several brain regions: particularly anterior cingulate cortex, frontal cortex, cortex, hippocampus, amygdala, and the basal ganglia structures caudate nucleus, putamen, and nucleus accumbens (Supplementary Figure 3). Using Brain RNA-Seq,^44^ we also investigated the cell type-specific expression in humans and found that all the mapped genes were significantly expressed in at least one type of brain cell investigated (Supplementary Figures 6-9). The mapped genes were not significantly associated with any gene set.

## Discussion

In this study, we utilized the largest available GWAS data on common epilepsies and COG to provide new insights into their genetic relationship. Using MiXeR, we estimated substantial genetic overlap between COG and the four epilepsy phenotypes ‘all epilepsy’, GGE, JME and CAE (Fig. 1), in which almost all epilepsy risk variants were found to also affect cognitive performance. Moreover, using conjFDR we identified 66 distinct genomic loci shared between epilepsies and COG (Fig. 2). Among these shared loci, five loci were novel for ‘all epilepsy’, 27 for GGE, five for CAE and five for JME (Table 2, Supplementary Tables 4-7). Taken together, our study extends previous work by estimating extensive genetic overlap between several epilepsy phenotypes and COG, and we dissect their shared genetic basis by identifying several novel shared loci.

**Figure 1.**
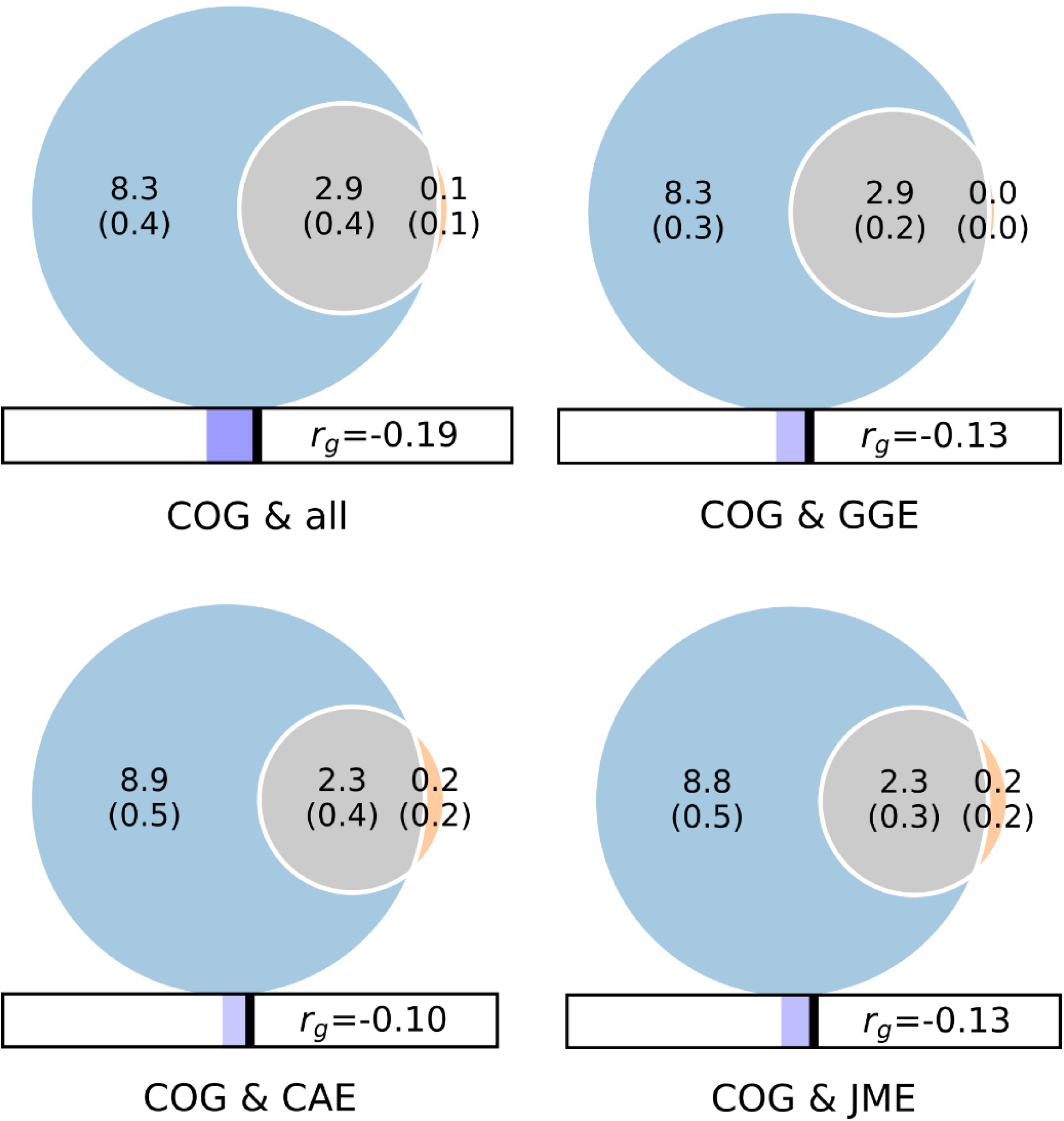
Genome-wide genetic overlap and genetic correlations between general cognitive ability (COG) and epilepsies (‘all epilepsy’, GGE, JME, CAE). The numbers in the Venn diagrams represent the number of shared and phenotype-specific trait-influencing variants which account for 90% of SNP-heritability in thousands, and *r*_g_ represents genome-wide genetic correlations. Abbreviations: COG, general cognitive ability; ALL, ‘all epilepsy’; GGE, genetic generalized epilepsies; JME, juvenile myoclonic epilepsy; CAE, childhood absence epilepsy)

**Figure 2.**
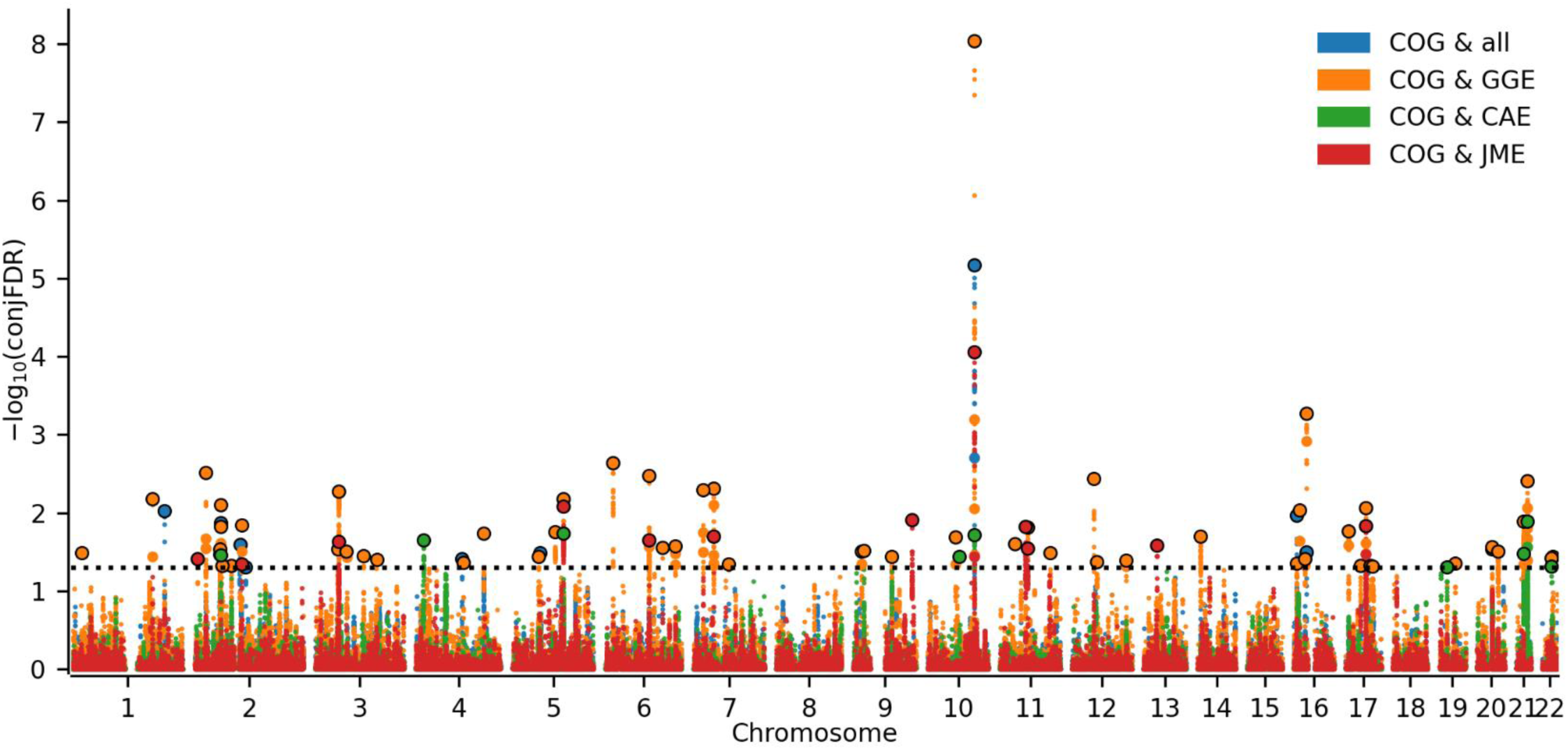
Common genetic variants jointly associated with COG and epilepsies (‘all epilepsy’, GGE, JME and CAE) at conjunctional false discovery rate (conjFDR) < 0.05. Manhattan plots showing the –log10 transformed conjFDR values for each single nucleotide polymorphism (SNP) on the y-axis and chromosomal positions along the x-axis. The dotted line represents the conjFDR threshold for significant association < 0.05. Abbreviations: COG, general cognitive ability; ALL, ‘all epilepsy’; GGE, genetic generalized epilepsies; JME, juvenile myoclonic epilepsy; CAE, childhood absence epilepsy)

**TABLE 2.**
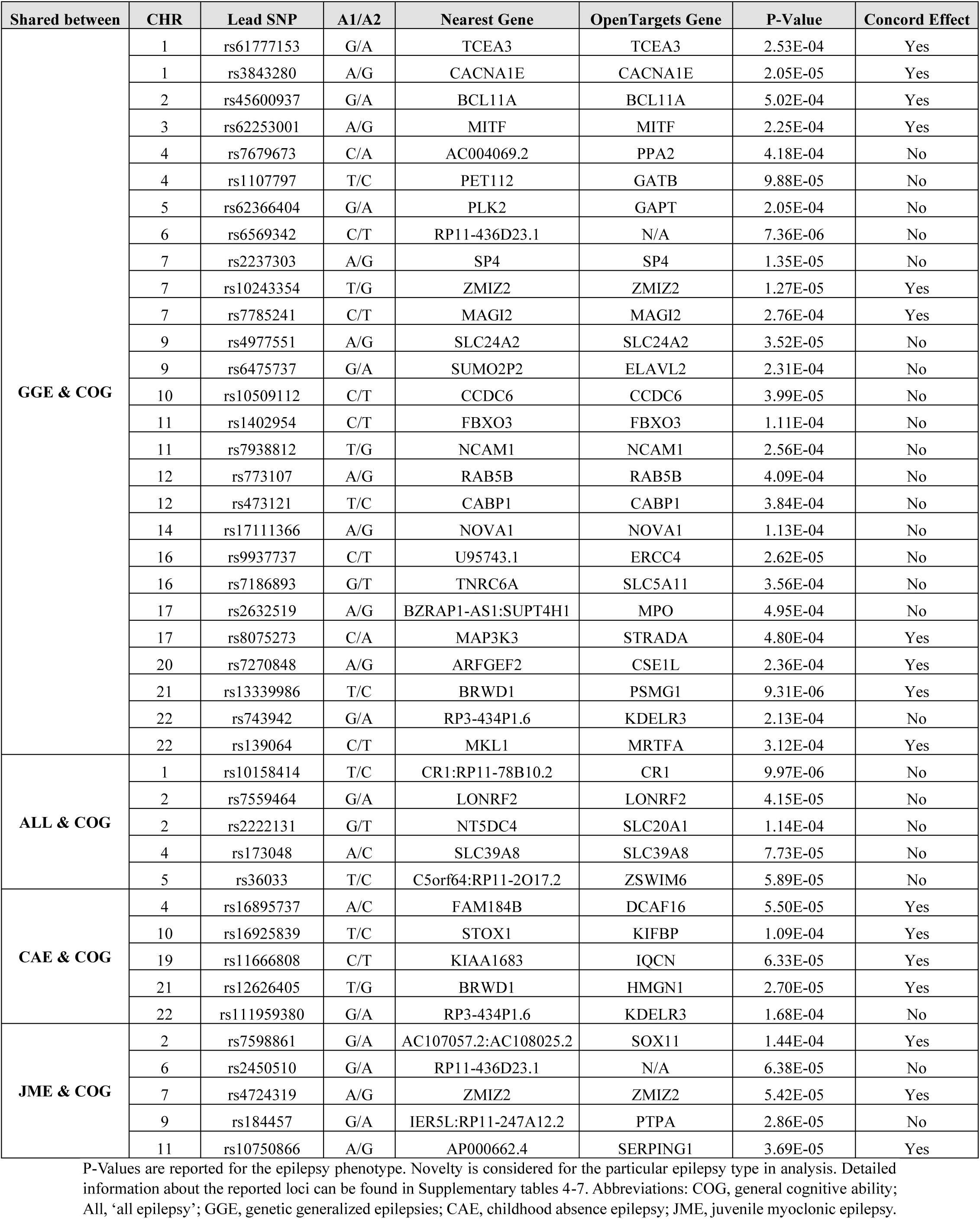
All novel loci associated with epilepsies at conjFDR<0.05.

Using MiXeR, we estimated COG to be almost four times more polygenic than the four epilepsy phenotypes ‘all epilepsy’, GGE, JME and CAE. As such, the estimated overlap represents a considerably smaller portion of the genetic architecture of COG than of the epilepsy phenotypes. While the majority of epilepsy risk variants were associated with lower cognitive performance, in line with negative correlations between these phenotypes,^16,17,21,22^ a large fraction of the epilepsy variants was associated with higher cognitive performance, indicating a complex genetic relationship between epilepsy and COG. These findings are consistent with the wide distribution of cognitive performance observed among individuals with epilepsy, as not all patients display poor cognitive functioning.^5,7^ Moreover, impairment in specific cognitive domains is differently associated with different epilepsy subgroups.^5^ As such, further studies are needed to investigate the genetic relationship of specific cognitive phenotypes and epilepsies. It would also be interesting to investigate the genetic link between COG and other predictive factors of severity of illness in epilepsy, including seizure onset, seizure frequency and duration, or comorbid psychiatric illness.^8^ For example, both epilepsy and COG are phenotypically and genetically linked to mental disorders.^18,21,23^ As such, patients with epilepsies with comorbid mental illness may constitute a subgroup at higher risk of worse cognitive functioning. In the future, combining multimodal diagnostics including imaging, EEG, blood-based biomarkers, and omics for epilepsies may aid in personalized risk estimation with prediction algorithms and may provide better treatment options with precision medicine.

The results emphasize the genetic heterogeneity across common epilepsies, as indicated by the considerably smaller SNP-heritability estimates of focal epilepsies (*h*^2^ range = 0.03 – 0.25) compared to the GGE types (*h*^2^ range = 0.59 – 0.95), which affected the ability to detect genetic overlap. The combination of relatively small GWAS samples and/or low SNP-heritability for the focal epilepsies leads to insufficiently powered MiXeR and conjFDR analyses. On the other hand, JME (*h*^2^ = 0.93, SE = 0.12) and CAE (*h*^2^ = 0.79, SE = 0.16) had larger SNP-heritability estimates than GGE (*h*^2^ = 0.60, SE = 0.05). Yet, the larger sample size for GGE yielded more power for conjFDR analyses and the greater part (83%) of the identified shared loci was linked to GGE. Accordingly, caution should be exercised when interpreting the present results. As the GWAS get larger, shared loci between several epilepsy types and COG are likely to be identified.

Among the overlapping loci, 15 loci were shared between at least two epilepsy types. A locus on chromosome 10 was significantly associated with all four epilepsy phenotypes and had the strongest association of all loci identified (top lead SNP rs11191116, conjFDR = 9.14 x 10^−9^ for COG and GGE), in which epilepsy risk was associated with lower cognitive performance. This locus reached genome-wide significance in all epilepsy phenotypes and COG.^17,19^ Our gene mapping approach implicated *KCNIP2* for this locus, which encodes a member of the voltage-gated potassium channel-interacting proteins (KCNIPs).^43^ This channel has recently been identified as a key regulator of homeostatic excitability in humans, and deletion leads to increased susceptibility to epilepsy and increased excitability in pyramidal hippocampal neurons.^48^ We also observed other mapped genes from the shared loci linked to membrane transport and signal transduction (*CACNA1E, CD47, SLC5A11, SLC24A2, CABP1, CPNE1*). In addition, many mapped genes are involved in transcription regulation (*SOX11, SOX14, HMGN1, TCEA3, ELL2, SP4*), neurodevelopmental processes (*KIFBP, SEMA3F, and NCAM1*), and serine/threonine kinase processes (*PTPA, VRK2* and *STRADA*).^43^ To our knowledge, 12 of these are novel gene associations for epilepsy (*CACNA1E*, *SLC5A11*, *SLC24A2*, *CABP1*, *SOX11*, *HMGN1*, *TCEA3*, *SP4*, *KIFBP*, *NCAM1*, *PTPA* and *STRADA*). In a previous study from our group identifying shared genomic loci between epilepsy and psychiatric disorders, we also observed several genes encoding serine/threonine kinases.^18^ Serine/threonine kinases are vital in the regulation of neuronal and synaptic activity, including the transport of neurotransmitters.^49^ Furthermore, among the 38 novel loci identified, six contained nonsynonymous exonic variants, within genes *LONRF2, FBXO3, TNRC6A, BRWD1, IER5L* and *SERPING1*. Nonsynonymous exonic variants are more likely to substantially affect a phenotype by disrupting protein function. The strongest novel association detected in our analysis was identified on chromosome 21 (top lead SNP rs13339986, conjFDR = 3.9 x 10^-3^ shared between GGE and COG), in which epilepsy risk was associated with higher cognitive performance. The locus was also shared with CAE, and included the nonsynonymous exonic variant within *BRWD1,* which encodes a member of the WD repeat family that regulates various cellular functions including signal transduction and cellular differentiation and plays a role in neurodevelopment. ^50^

The shared genetic signal between traits may, however, reflect both shared or separate causal variants in strong LD with each other, which conjFDR analysis cannot distinguish.^27^ As such, further experimental validation is required to determine how the identified genetic variants impact cognitive performance and the risk of epilepsies. Nevertheless, with functional annotation, we marked several candidate SNPs that may be plausible causal variants in the shared loci for follow-up studies. Another limitation was the use of European ancestry datasets to avoid LD bias in conjFDR analyses. This means that our results may not be translatable to other ancestries at this time. This limitation can be overcome with larger GWAS samples for epilepsies and COG based on other ancestries.

## Conclusion

Overall, we demonstrate polygenic overlap between several epilepsy types and COG with significant negative correlations. In addition, we discover multiple novel shared genomic loci between these phenotypes. In summary, the present study indicates that the pathogenic processes underlying epilepsies to a large extent also influence the biological underpinnings of COG.

## Data availability statement

GWAS summary statistics used in this study are publicly available and can be accessed through the original publications.

- MiXeR (github.com/precimed/mixer/commit/f56a44, https://github.com/comorment/mixer/tree/v1.3.0)
- CondFDR & ConjFDR (https://github.com/precimed/pleiofdr/commit/846441)
- LDSC (https://github.com/bulik/ldsc, github.com/comorment/ldsc/commit/f2d7d6)
- FUMA (https://fuma.ctglab.nl/)
- OpenTargets Genetics (https://genetics.opentargets.org/)
- Cleansumstats (https://github.com/BioPsyk/cleansumstats/tree/1.6.0)
- Brain RNA-Seq (https://brainrnaseq.org/)
- MATLAB R2017a (https://se.mathworks.com)

## Author Contributions

Conceived and designed the analysis: N.K., E.H., O.A.A., O.B.S.; Contributed data or analysis tools: E.H., A.A.S., Z.R., G.K., N.P., O.F., A.M.D., O.A.A. ; Performed the analysis: N.K., E.H., A.A.S., G.K., N.P.; Drafting the article: N.K., E.H., O.B.S.; Critical revision of the article: N.K., E.H., A.A.S., D.V.D.M., K.S.O., Z.R., G.K., N.P., S.B., V.F., K.H., E.T., T.U., N.E.S., S.D., A.M.D., O.F., O.A.A., O.B.S.; Final approval of the version to be published: N.K., E.H., A.A.S., D.V.D.M., K.S.O., Z.R., G.K., N.P., S.B., V.F., K.H., E.T., T.U., N.E.S., S.D., A.M.D., O.F., O.A.A., O.B.S.

## Data Availability

https://github.com/precimed/mixer/commit/f56a44

https://github.com/comorment/mixer/tree/v1.3.0

https://github.com/precimed/pleiofdr/commit/846441

https://github.com/bulik/ldsc

https://github.com/comorment/ldsc/commit/f2d7d6

https://fuma.ctglab.nl/

https://genetics.opentargets.org/

https://github.com/BioPsyk/cleansumstats/tree/1.6.0

https://brainrnaseq.org/

https://se.mathworks.com

## Acknowledgements

All GWAS investigated in the present study were approved by the local ethics committees, and informed consent was obtained from all participants. We thank the ILAE consortium and the FinnGen biobank for access to data, and the many participants who provided DNA samples.

This work was performed on Services for sensitive data (TSD), University of Oslo, Norway, with resources provided by UNINETT Sigma2 - the National Infrastructure for High Performance Computing and Data Storage in Norway.

The project is funded by ERA-NET-NEURON (Application no: ES609126, Project NMDAR- PSY) which is funded by participating Research Councils across Europe. The work was supported by the Research Council of Norway (262656, 249711, 248980, 248778, 226971, 223273, 300309, 324252, 324499, 326813 and 334920), South-East Norway Regional Health

Authority (2016–064, 2022-073 and 2022-087), KG Jebsen Stiftelsen (SKGJ-MED-008), European Union’s Horizon 2020 Research and Innovation Programme (847776, CoMorMent; 964874, RealMent; and 801133, Marie Sklodowska-Curie Actions) and EAA grant (#EEA-RO- NO-2018-0573). K.S.O., G.K. and A.M.D. were supported by NIH (K.S.O: 5R01MH124839- 02; G.K.: R01MH123724-01; A.M.D.: U24DA041123, R01AG076838, U24DA055330, and OT2 HL161847). S.B. was supported by Norwegian Health Association (22731).

## Disclosure of Conflicts of Interest

O.A.A. has received speaker’s honorarium from Lundbeck, Sunovion, Takeda, Janssen and is a consultant for CorTechs.ai and Precision Health AS. A.M.D. is a founder of and holds equity interest in CorTechs Labs and serves on its scientific advisory board. He is also a member of the Scientific Advisory Board of Healthlytix and receives research funding from General Electric Healthcare (GEHC). The terms of these arrangements have been reviewed and approved by the University of California, San Diego in accordance with its conflict-of-interest policies. Remaining authors have no conflicts of interest to declare.

## Notes

### Author Declarations

GWAS summary statistics used in this study are publicly available and can be accessed through the original publications. Each sample was collected with the participants written informed consent and with approval by local institutional review boards. The use of summary statistics for genetic analysis was evaluated by The Norwegian Institutional Review Board: Regional Committees for Medical and Health Research Ethics (REC) South-East Norway and found that no additional ethical approval was required because no individual data were used.

