## Supplementary Information for "Unraveling the shared genetics of common epilepsies and general cognitive ability"

\# Shared first-author

**Corresponding Author:** Olav B. Smeland

Centre for Precision Psychiatry, Division of Mental Health and Addiction

University of Oslo and Oslo University Hospital

Kirkeveien 166, 0424 Oslo, Norway

**Supplementary Information**

**Supplementary Figure 1**
Polygenic overlap between epilepsies and general cognitive ability.

**Supplementary Figure 2**
Cross-trait enrichment between epilepsies and general cognitive ability.

**Supplementary Figure 3**
Tissue enrichment for differential gene expression (DEG) in 54 GTEx tissue types for the genes mapped from the shared loci between epilepsies and general cognitive ability.

**Supplementary Figure 4**

Gene expression heatmap of the genes mapped from the shared loci between epilepsies and general cognitive ability across 54 human tissue types in the GTEx v8 dataset.

**Supplementary Figure 5**

Gene set enrichment of the mapped genes in the GWAS Catalogue.

**Supplementary Figure 6**

Brain expression quantitative trait loci investigation (Brain RNA-Seq) of the genes mapped from the shared loci between ‘all epilepsy’ and general cognitive ability.

**Supplementary Figure 7**

Brain expression quantitative trait loci investigation (Brain RNA-Seq) of the genes mapped from the shared loci between genetic generalized epilepsies and general cognitive ability.

**Supplementary Figure 8**

Brain expression quantitative trait loci investigation (Brain RNA-Seq) of the genes mapped from the shared loci between childhood absence epilepsy and general cognitive ability.

**Supplementary Figure 9**

Brain expression quantitative trait loci investigation (Brain RNA-Seq) of the genes mapped from the shared loci between juvenile myoclonic epilepsy and general cognitive ability.

**Supplementary Table A**

Overview of the mapped genes from the novel epilepsy loci.

**Supplementary Fig 1.**

**Polygenic overlap between epilepsies and general cognitive ability.**

Bivariate MiXeR analysis generates conditional quantile-quantile plots to visualize the cross-trait SNP enrichment (left and middle columns) and negative log-likelihood plots to indicate the performance of the best model vs. min and max (right column) for each pair of traits. Abbreviations: COG, general cognitive ability; All, ‘all epilepsy’; GGE, genetic generalized epilepsies; CAE, childhood absence epilepsy; JME, juvenile myoclonic epilepsy.

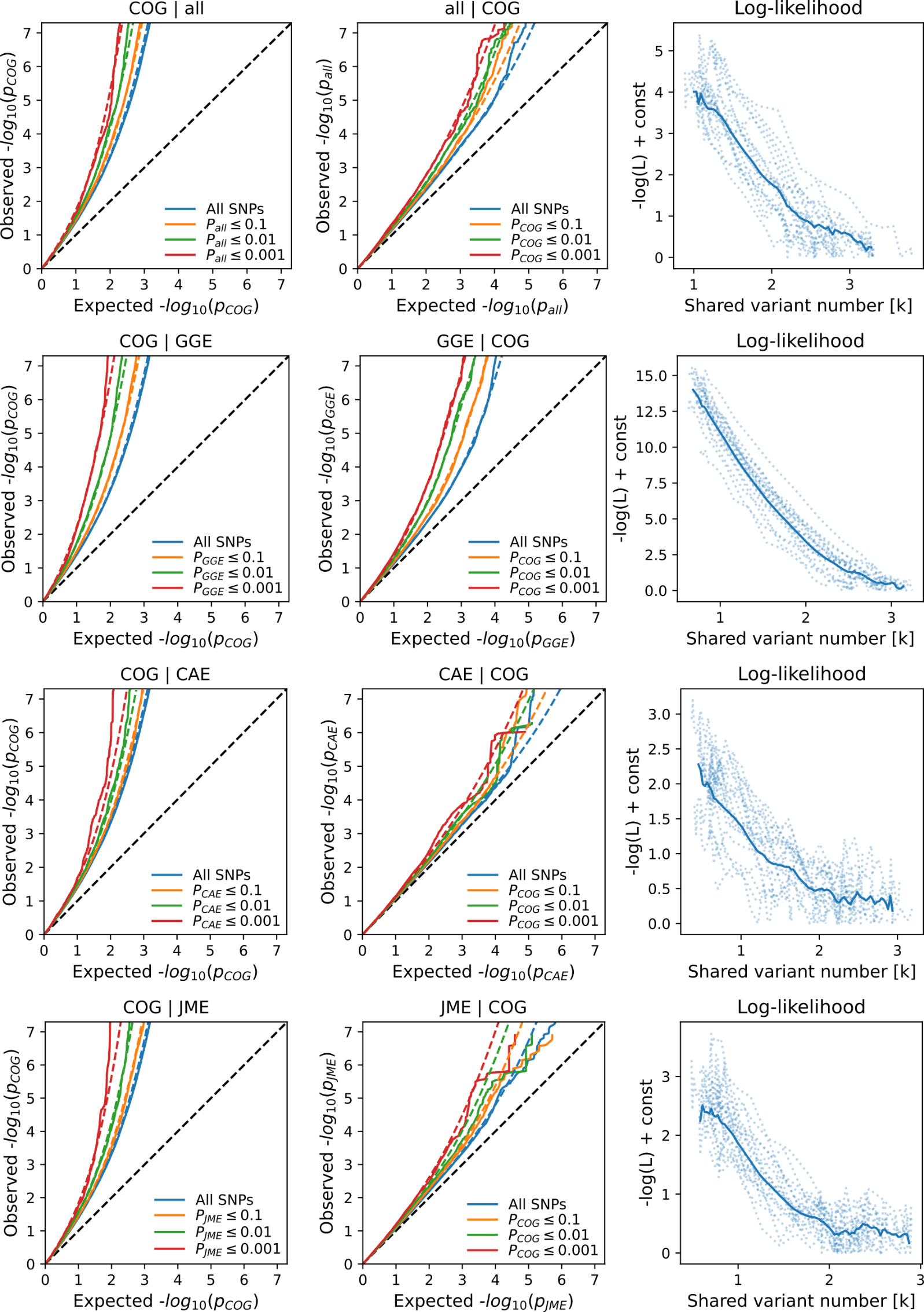

**Supplementary Fig 2.**

**Cross-trait enrichment between epilepsies and general cognitive ability.**

Quantile-quantile (Q-Q) plots showing single nucleotide polymorphism (SNP) enrichment for general cognitive ability conditional on SNP-associations with epilepsies (left column). Reverse Q-Q plots show SNP-enrichment for epilepsies (right column) conditional on SNP-associations with general cognitive ability. Conditional quantile-quantile plots of nominal versus empirical −log_10_(*p*) values (corrected for inflation) in primary trait below the standard genome-wide association study threshold of *p* < 5 × 10^−8^ as a function of significance of association with the secondary trait, at the level of *p* < 0.10, *p* < 0.01 and *p* < 0.001. The blue lines indicate all single nucleotide polymorphisms (SNPs). The dashed lines indicate the null hypothesis. Abbreviations: COG, general cognitive ability; All, ‘all epilepsy’; GGE, genetic generalized epilepsies; CAE, childhood absence epilepsy; JME, juvenile myoclonic epilepsy.

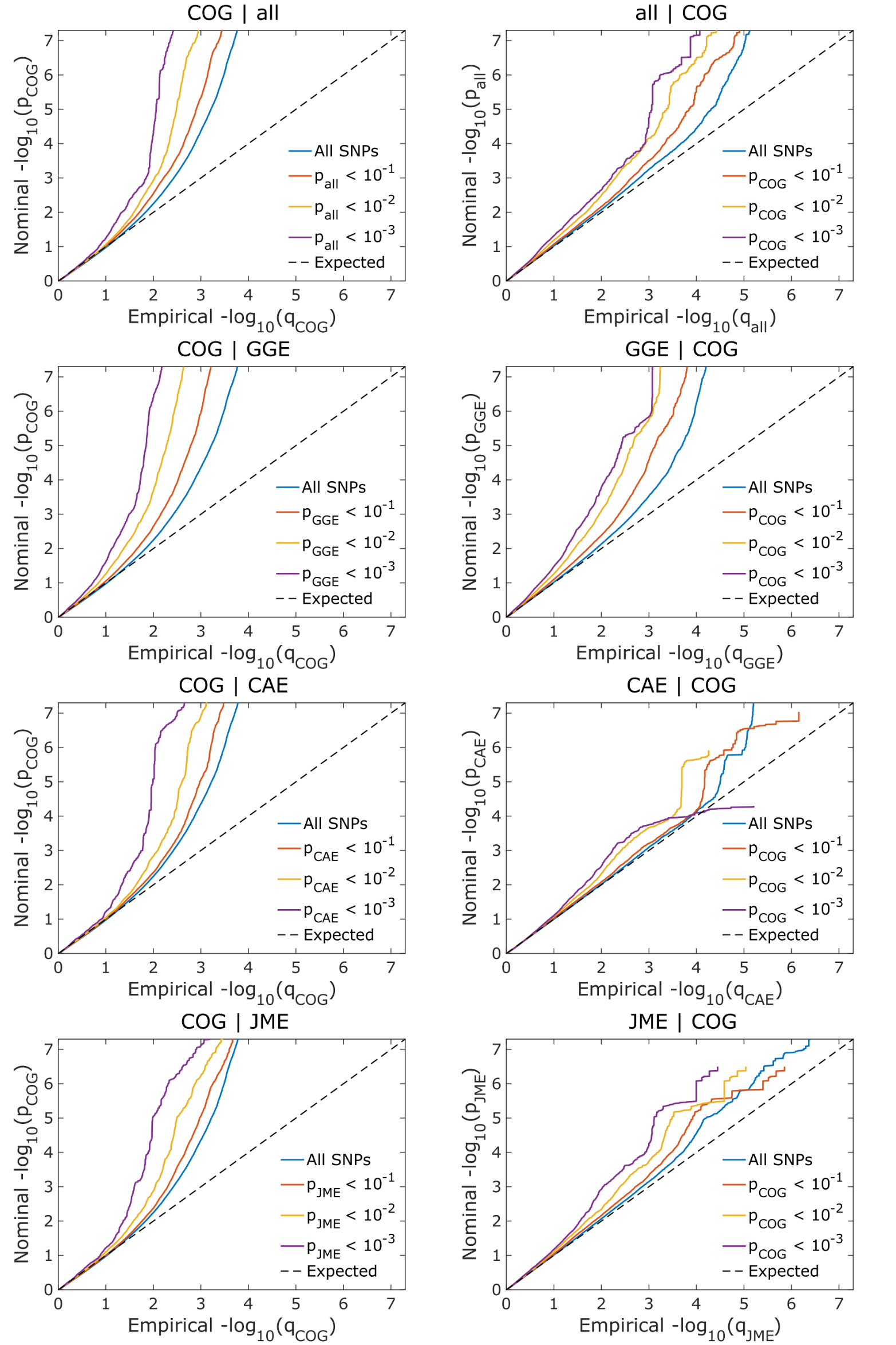

**Supplementary Fig 3.**

**Tissue enrichment for differential gene expression (DEG) in 54 GTEx tissue types for the genes mapped from the shared loci between epilepsies and general cognitive ability.**

DEG sets significantly enriched with the prioritized genes are highlighted in red, shown for higher (up-regulated DEG), lower (down-regulated DEG), or two-sided differences in gene expression [DEG (both sides)]. The analysis was performed using FUMA.

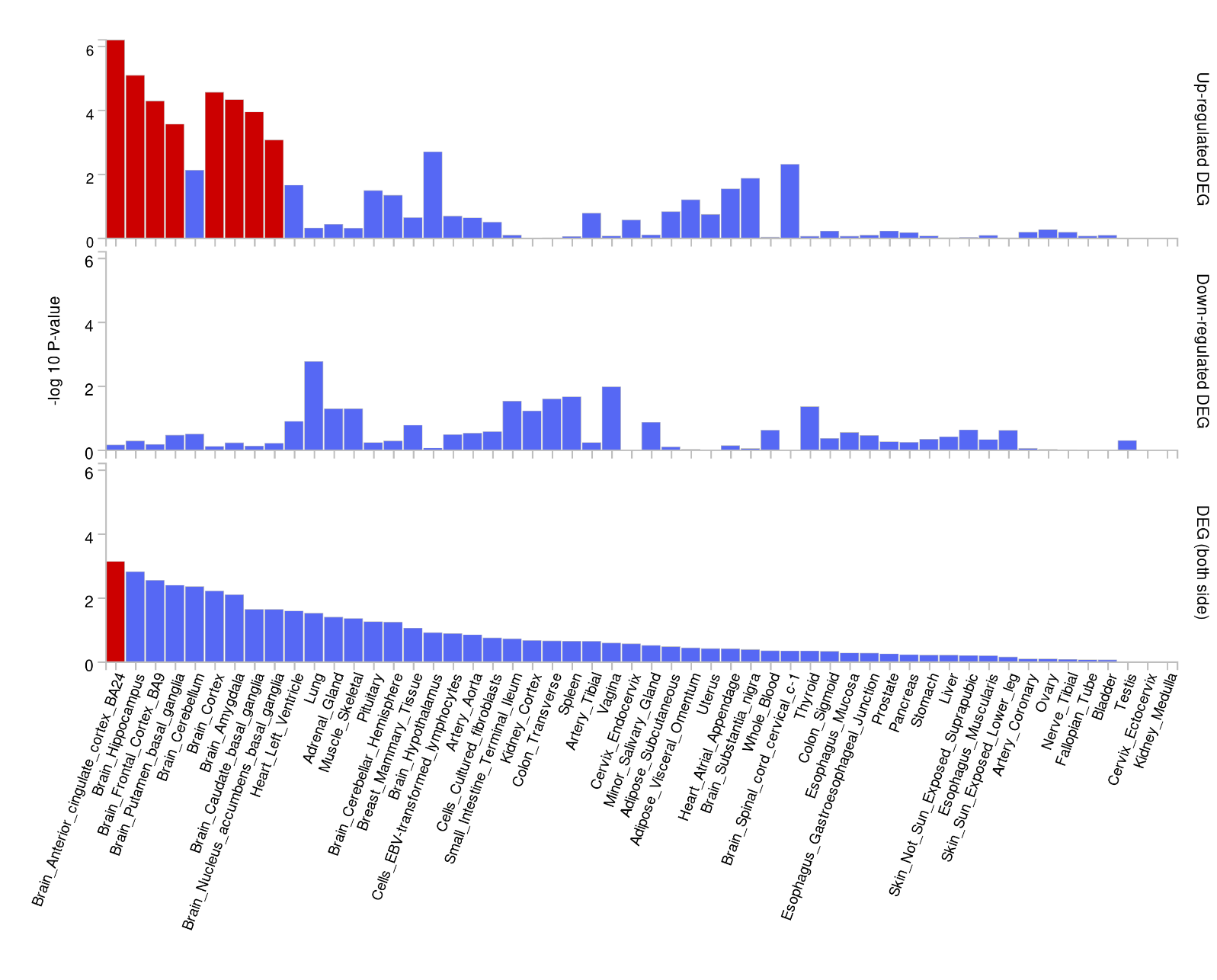

**Supplementary Fig 4.**

**Gene expression heatmap of the genes mapped from the shared loci between epilepsies and general cognitive ability across 54 human tissue types in the GTEx v8 dataset.**

Expression values are shown as an average of log_2_ transformed per-label values in the GTEx v8 dataset, as implemented in FUMA. Darker red colors represent higher gene expression compared to darker blue colors. Mapped genes are shown on the x-axis while the tissue types are shown on the y-axis. The genes and the tissues are ordered by cluster.

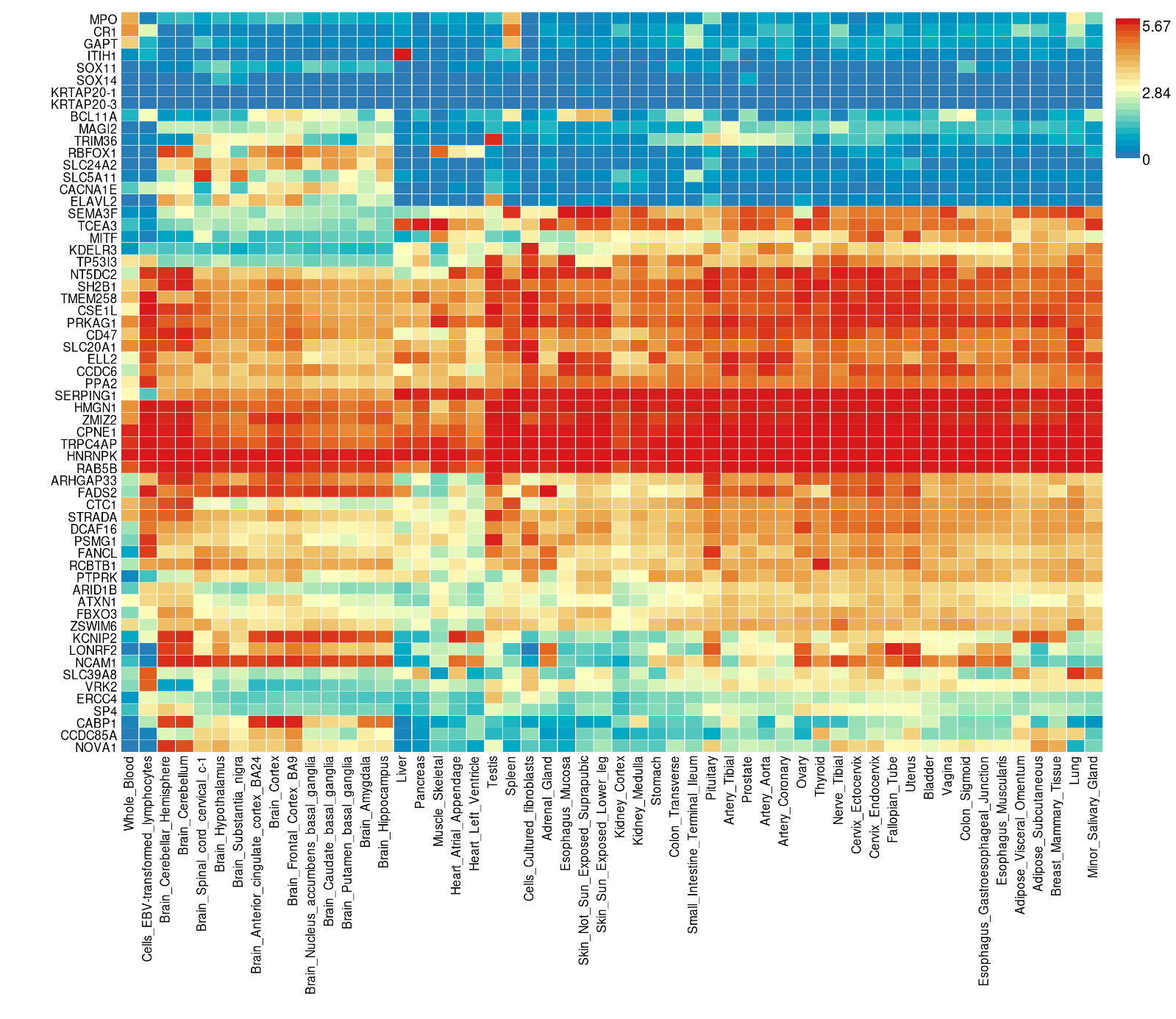

**Supplementary Figure 5.** **Gene set enrichment of the mapped genes in GWAS Catalogue.**

The association between the mapped genes from the identified shared loci and gene sets as defined by their phenotype association in the GWAS catalogue.^1^ Only significant gene sets are shown after correcting for multiple comparisons using FDR correction.

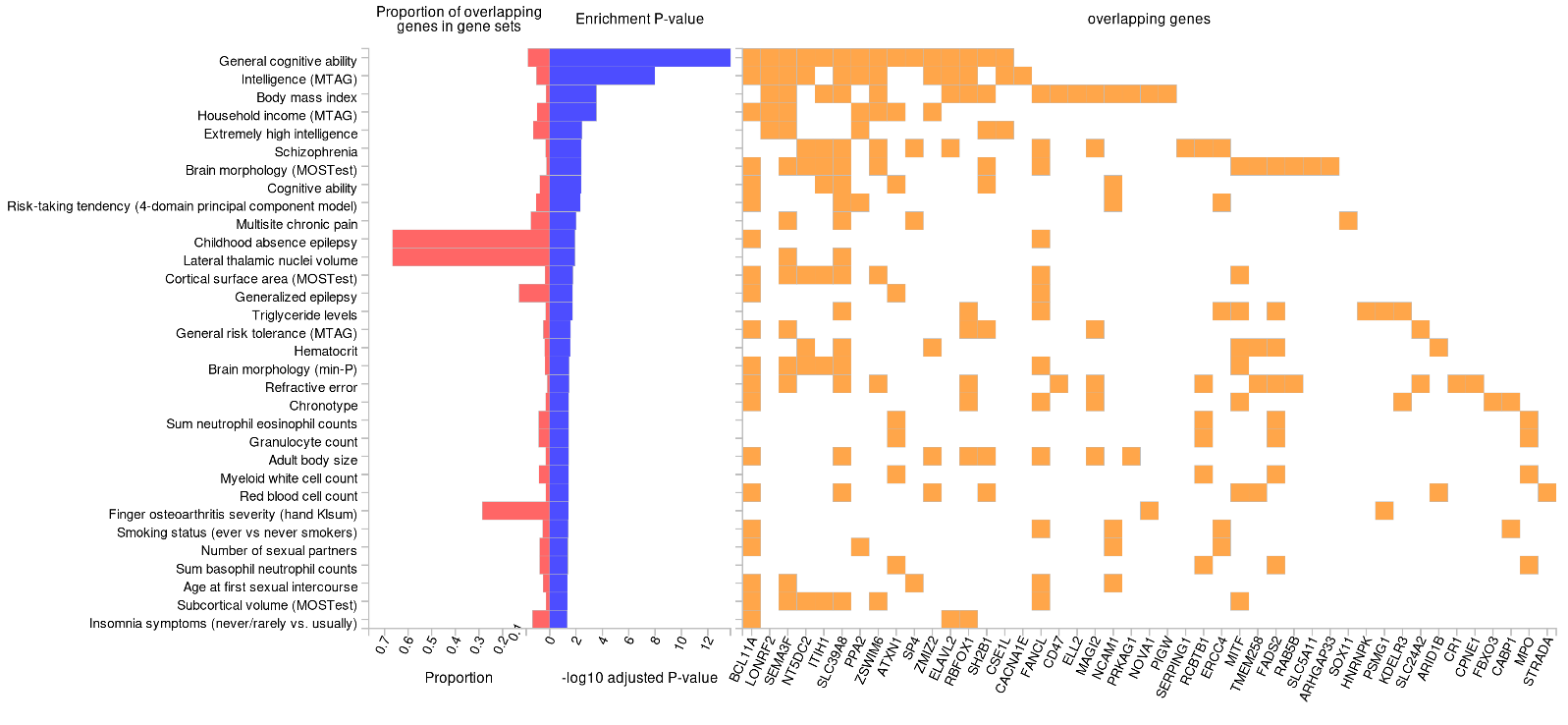

**Supplementary Figure 6**

Single-cell RNA sequencing investigation (Brain RNA-Seq) of the genes mapped from the shared loci between ‘all epilepsy’ and general cognitive ability shows that the mapped genes are significantly expressed in brain cells.

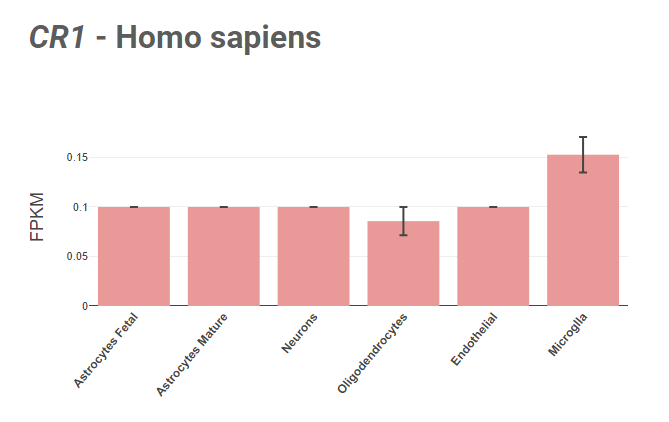

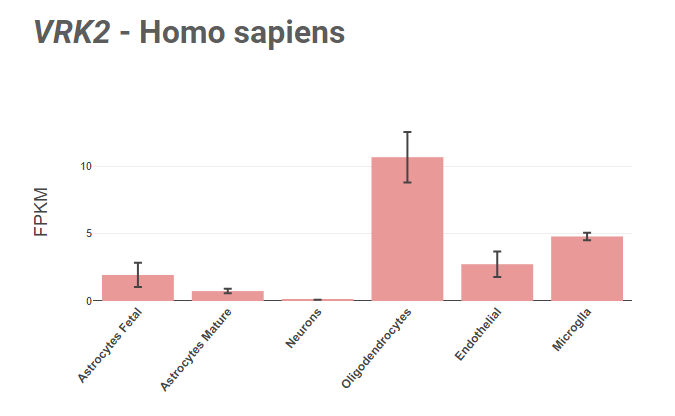

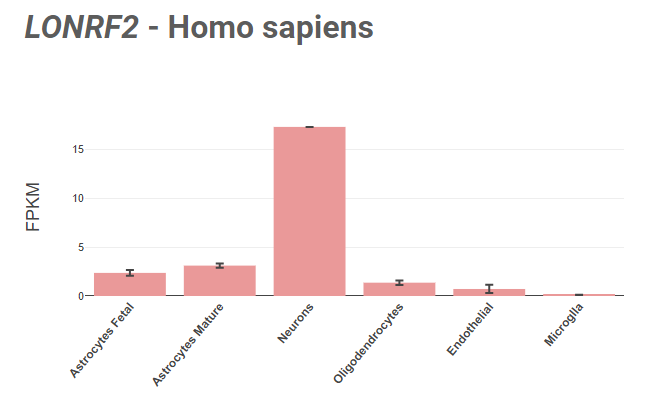

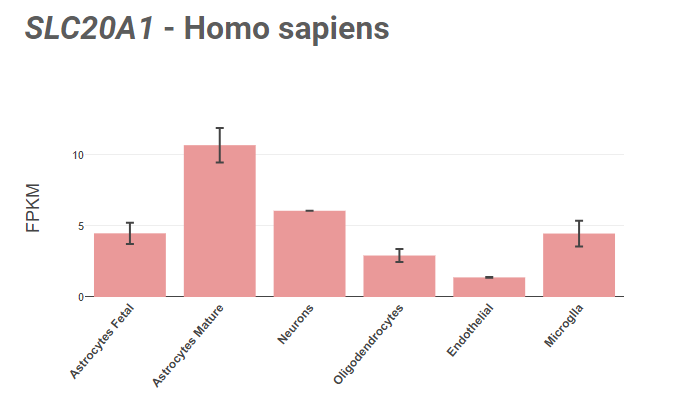

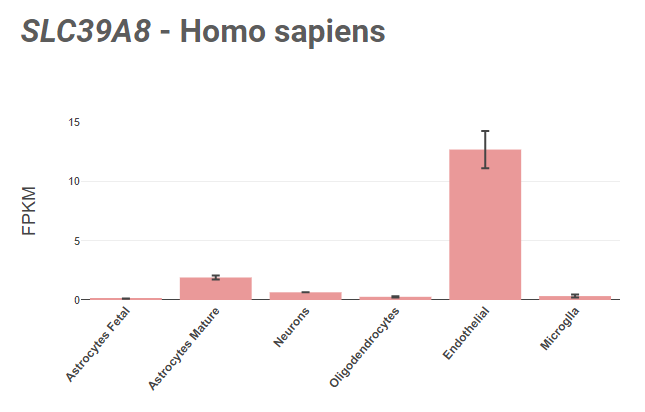

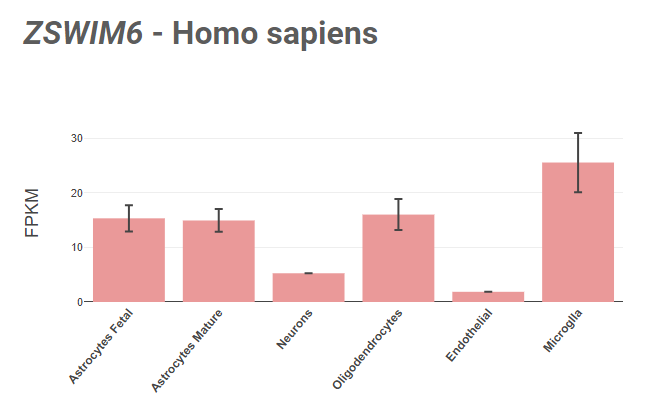

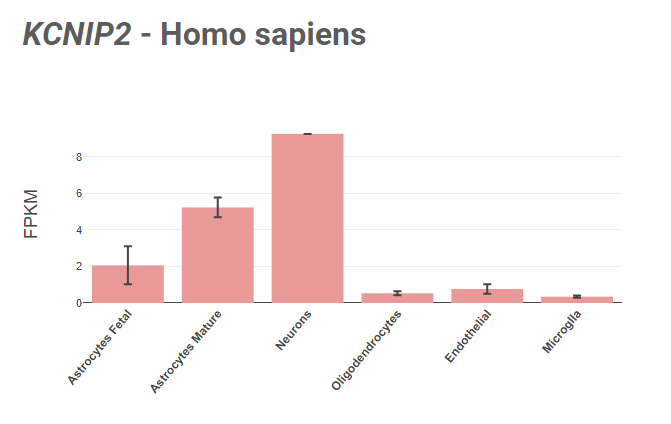

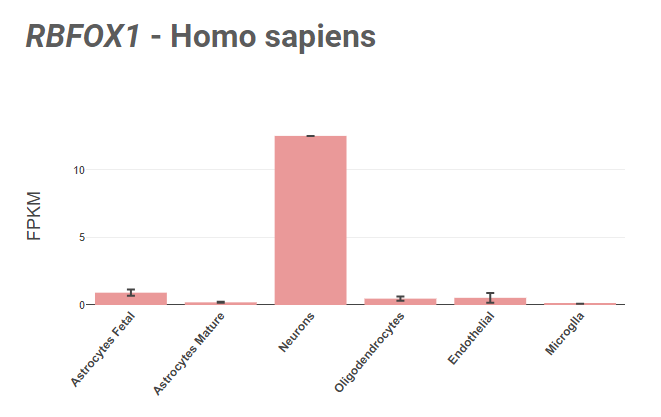

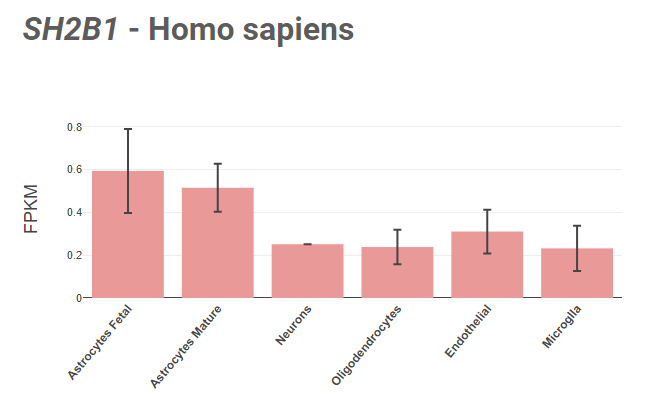

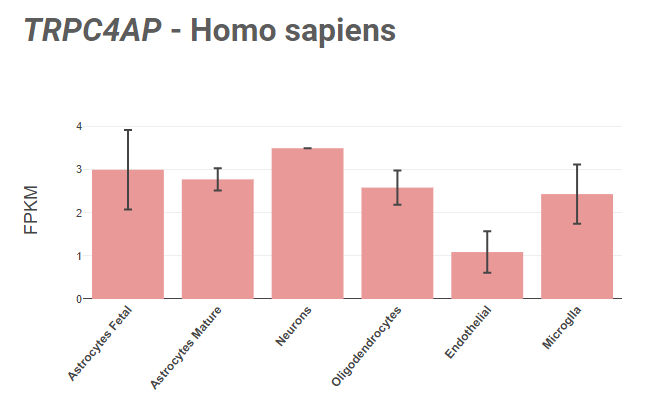

**Supplementary Figure 7**

Single-cell RNA sequencing investigation (Brain RNA-Seq) of the genes mapped from the shared loci between genetic generalized epilepsies and general cognitive ability shows that the mapped genes are significantly expressed in brain cells.

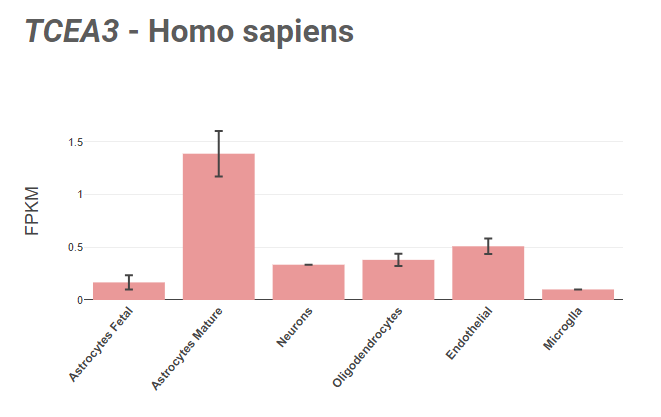

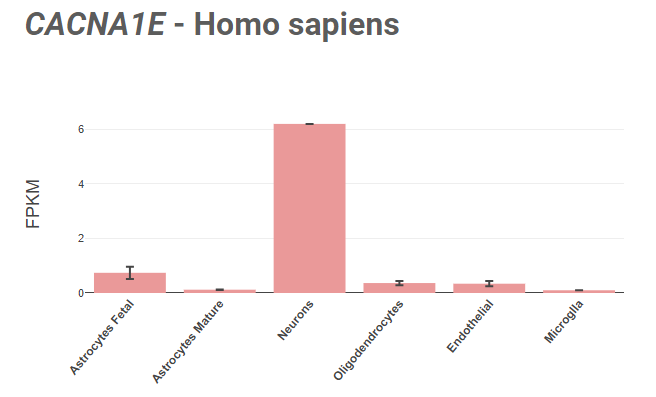

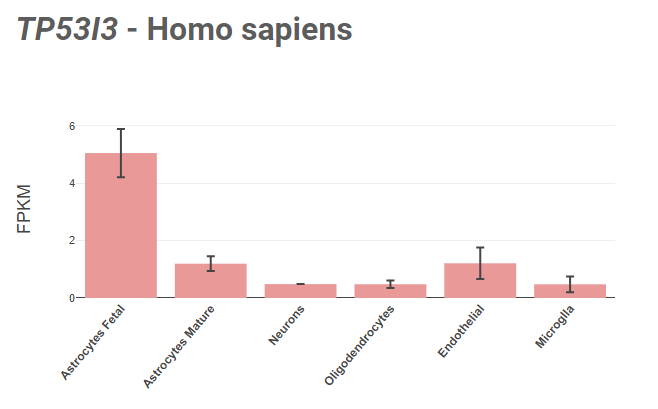

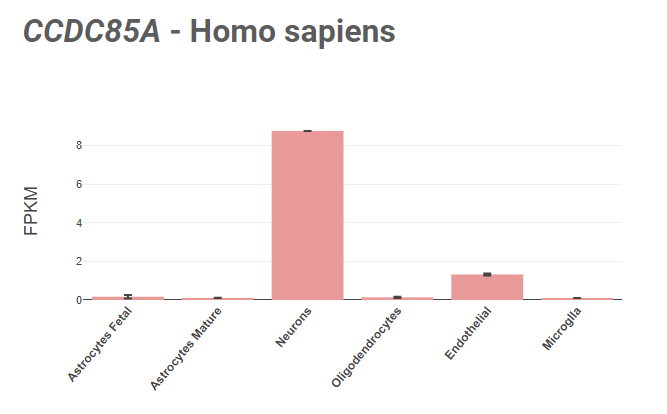

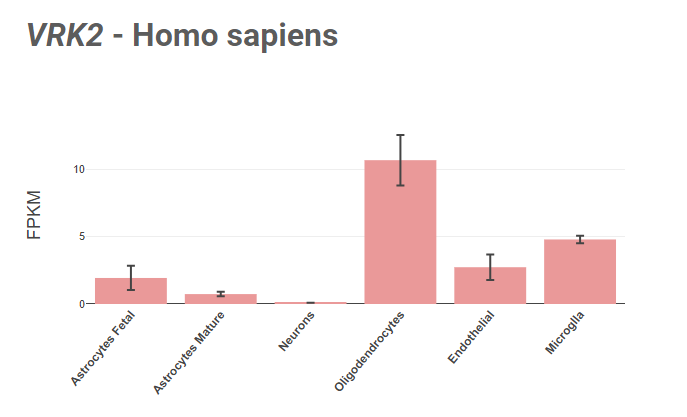

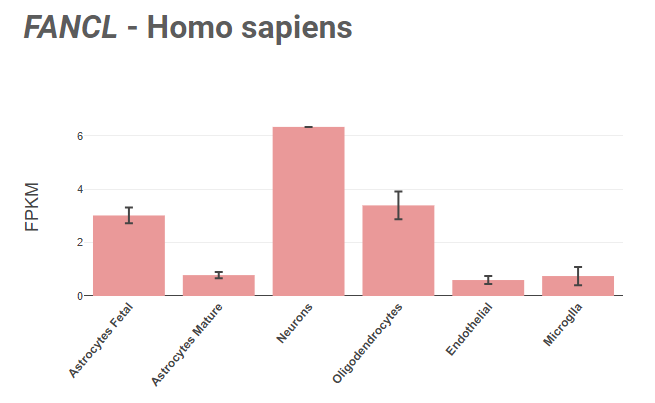

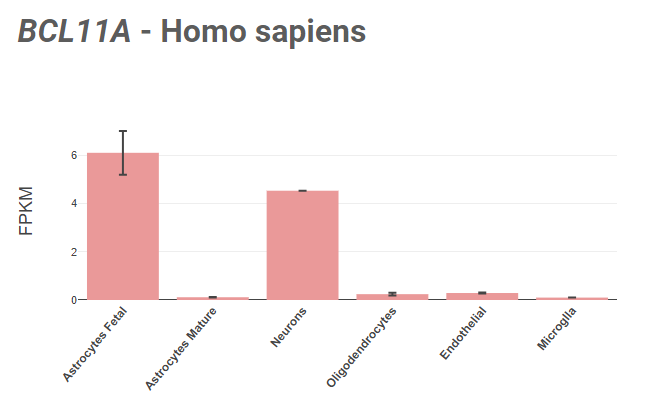

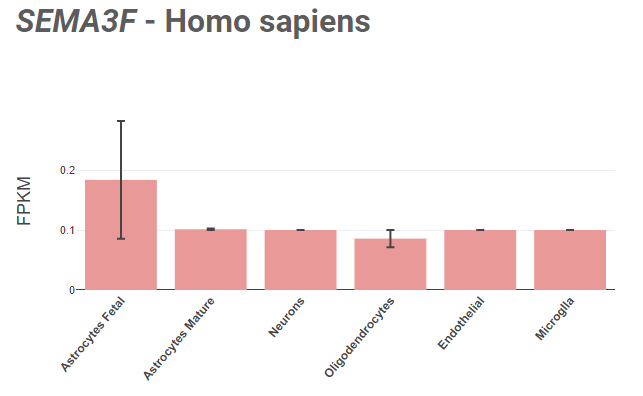

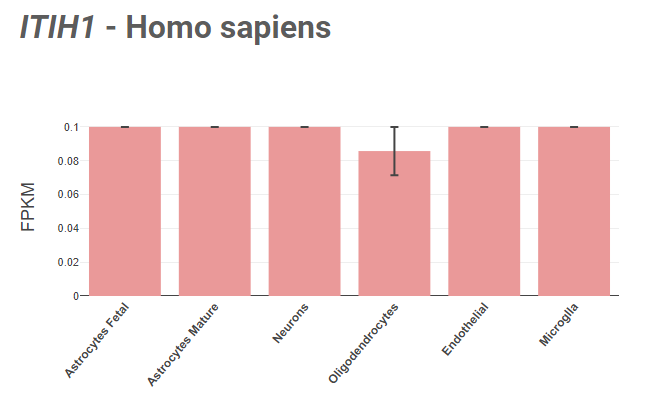

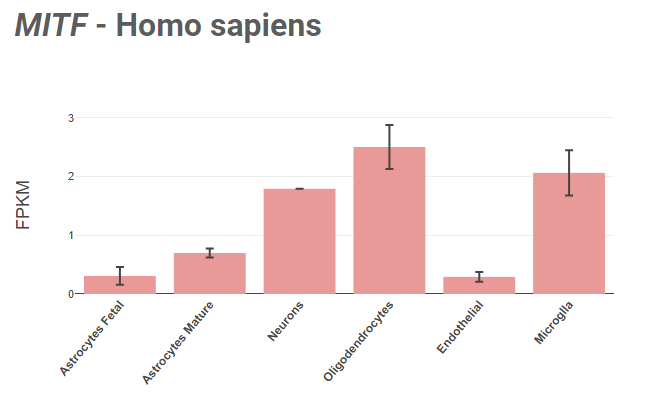

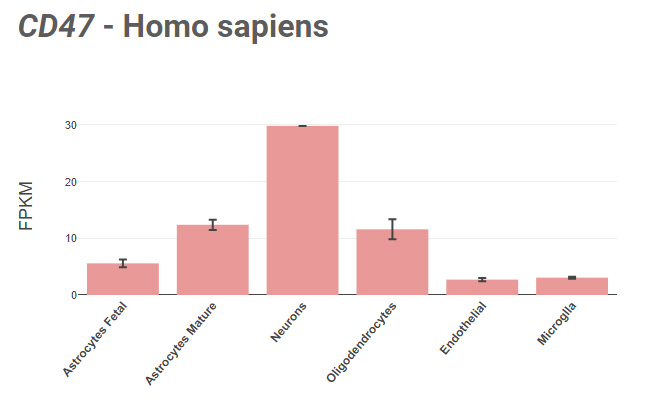

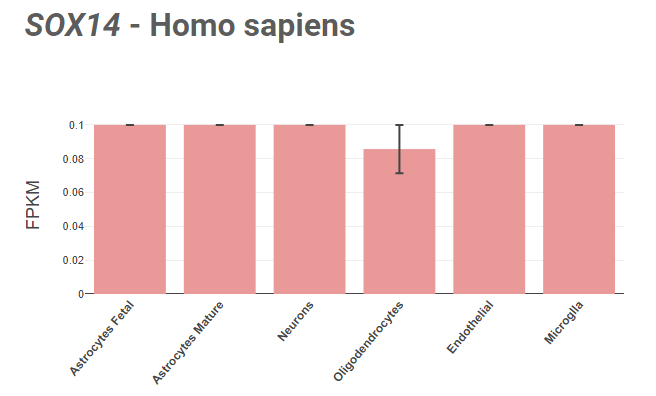

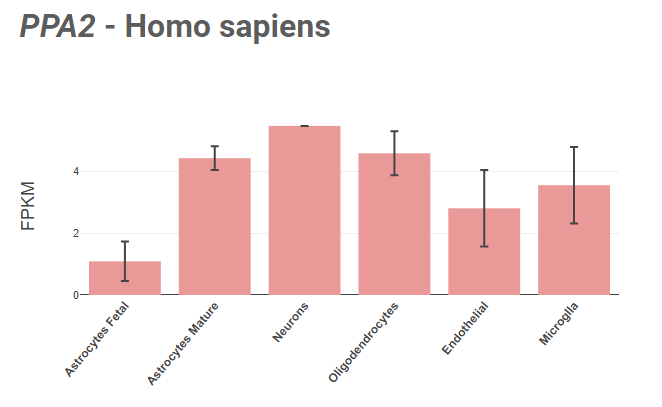

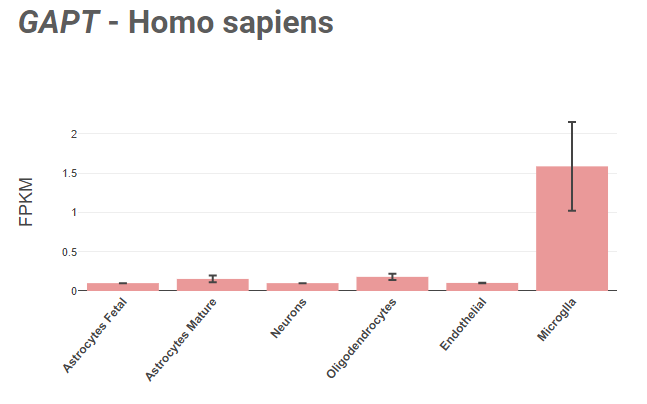

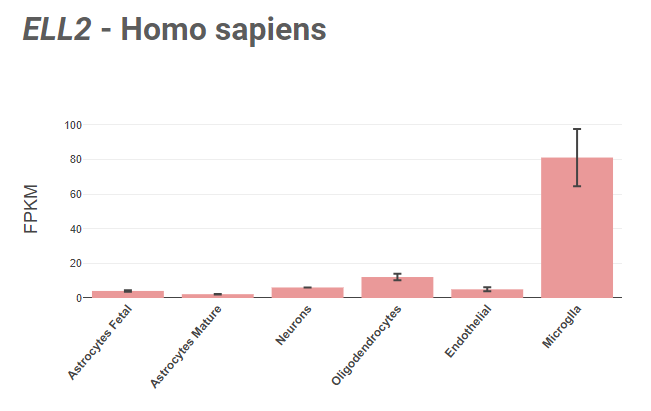

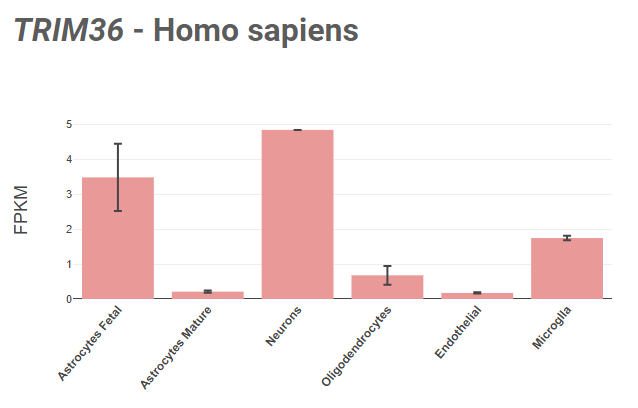

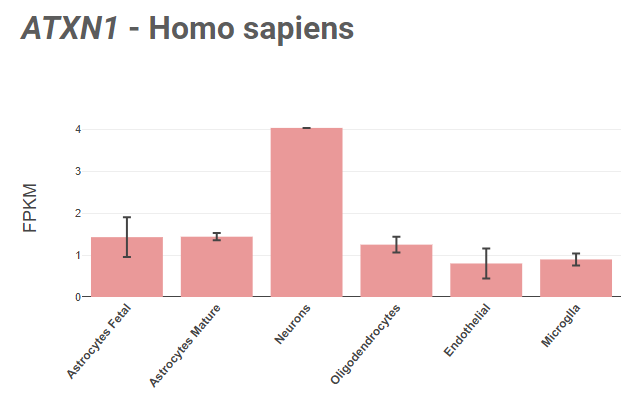

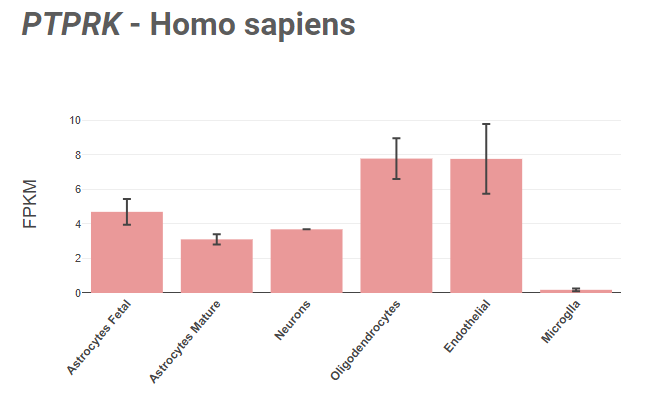

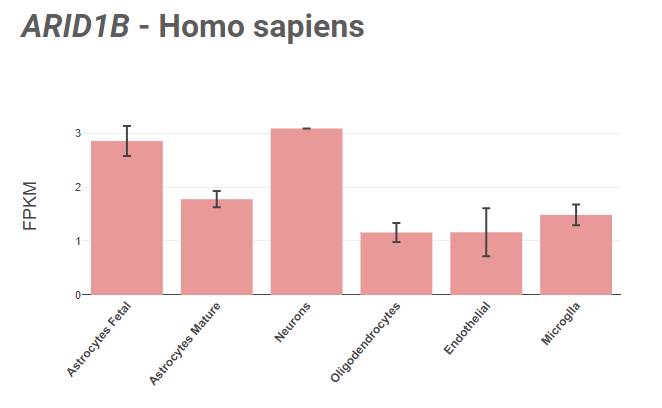

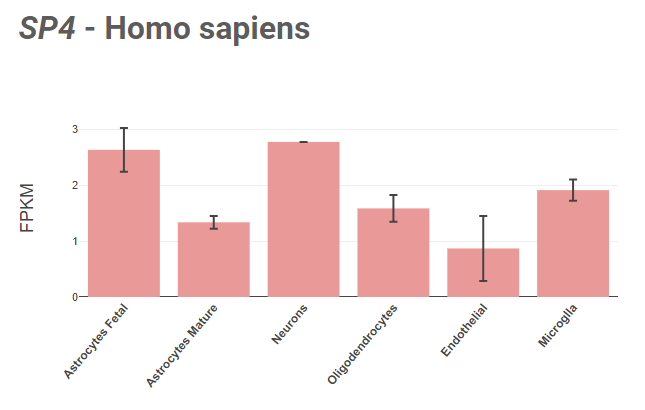

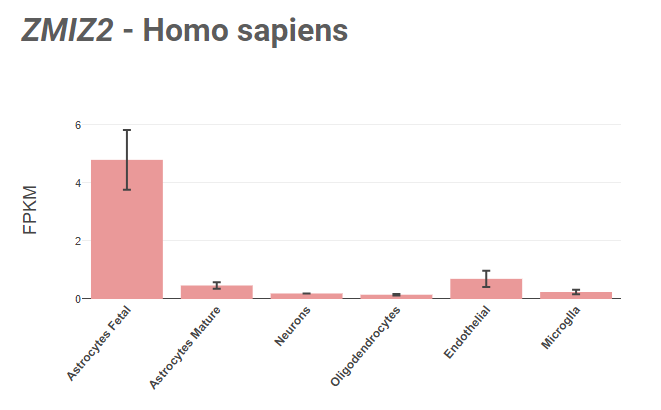

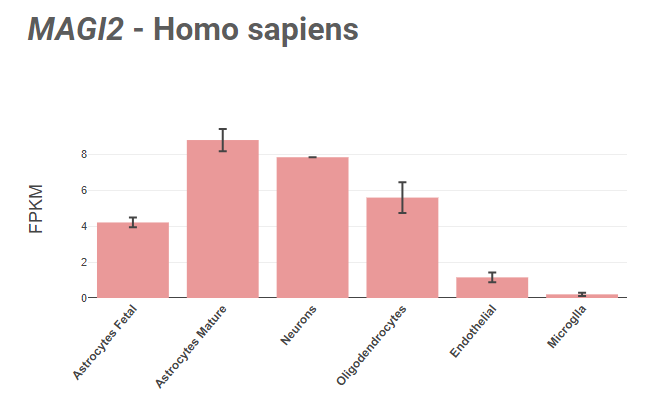

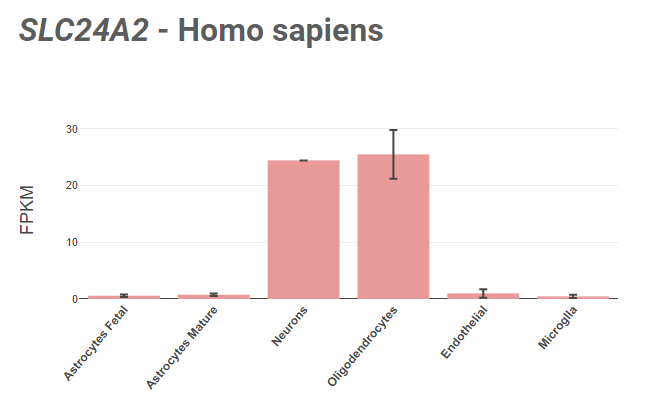

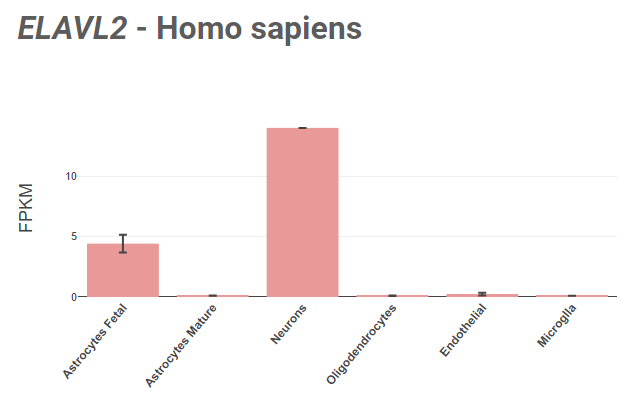

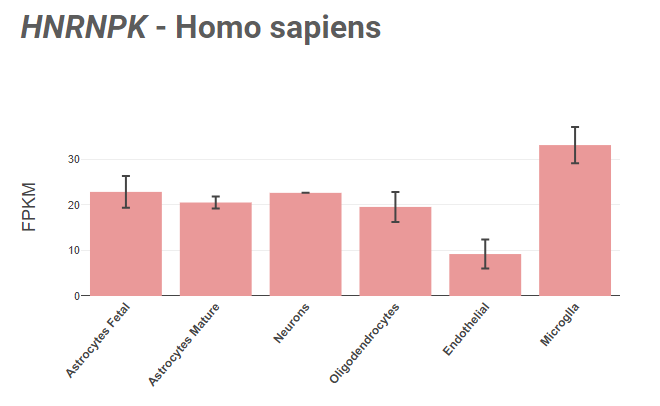

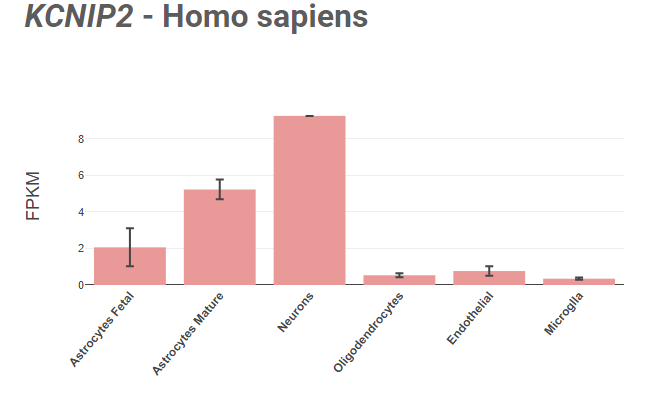

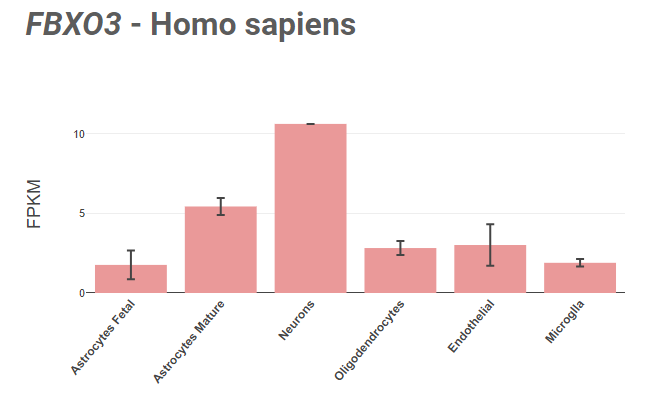

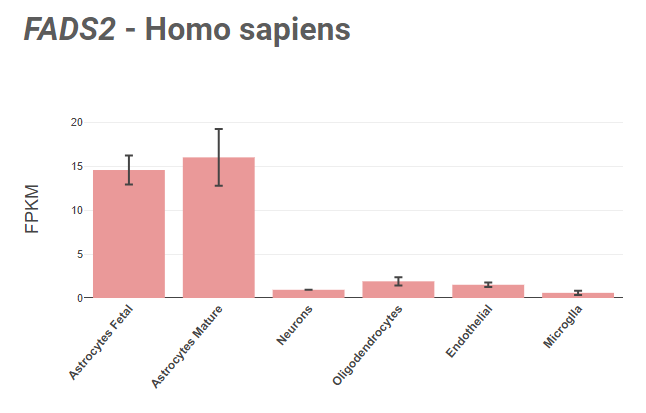

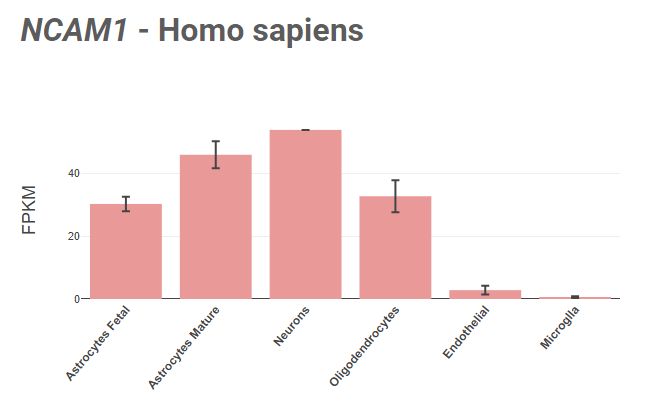

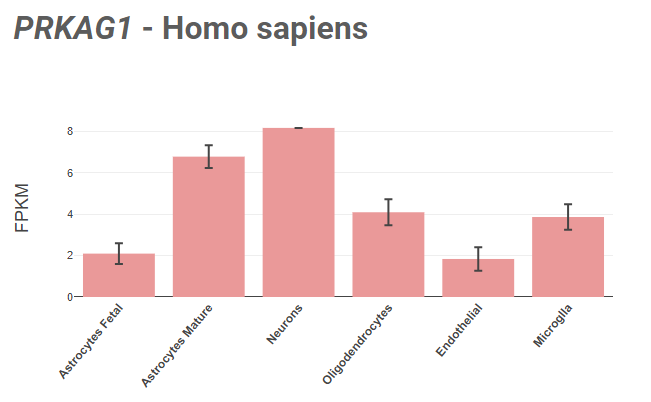

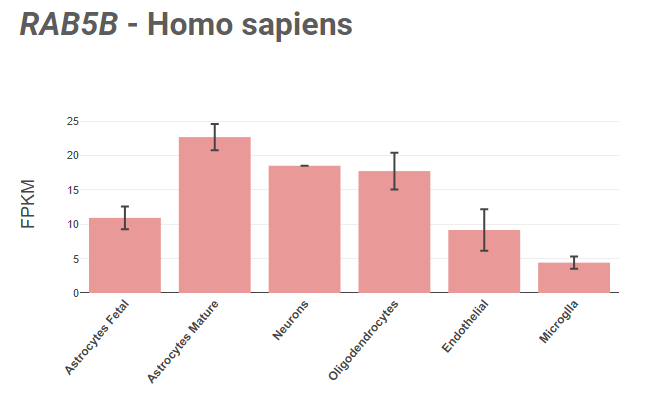

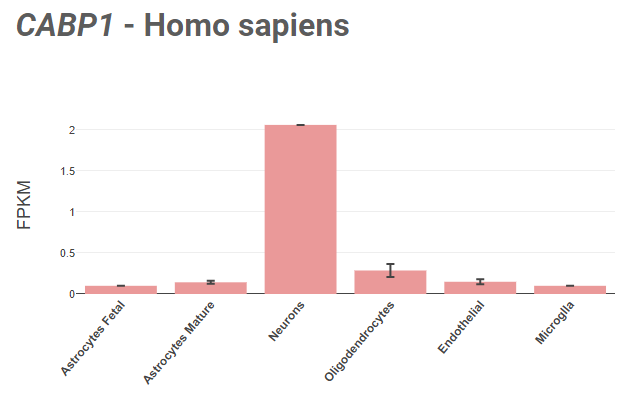

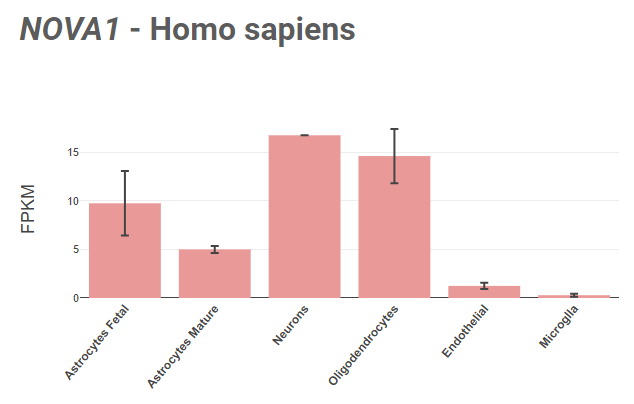

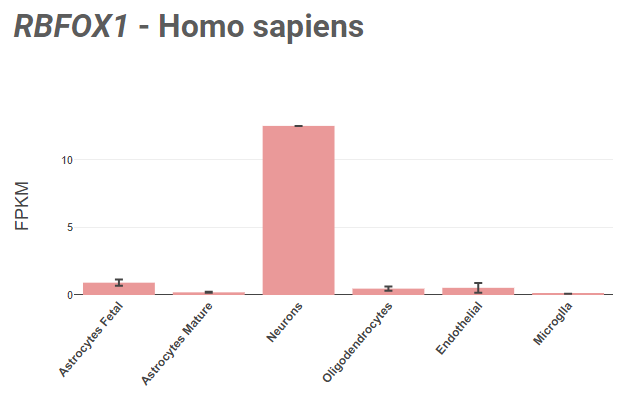

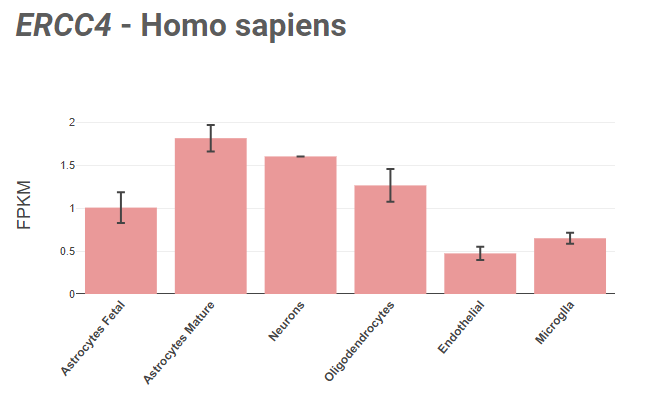

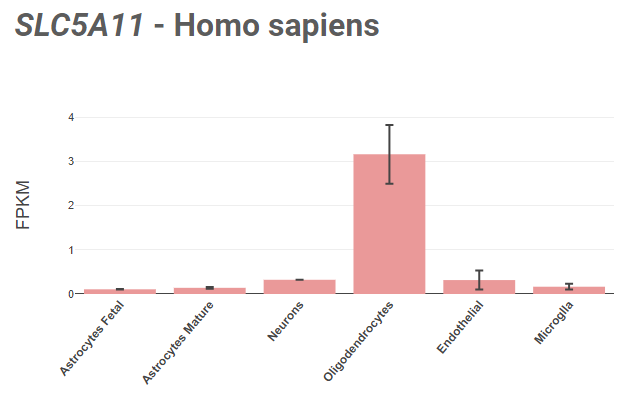

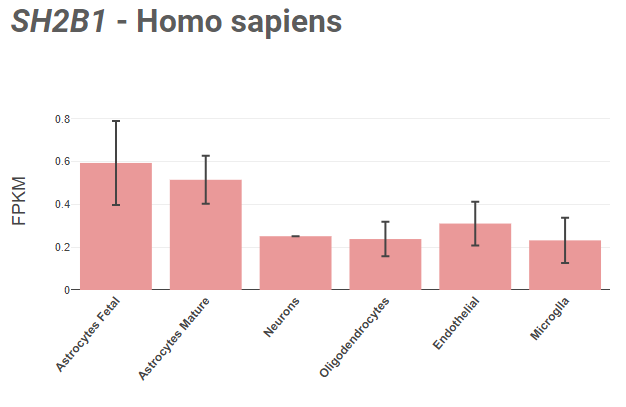

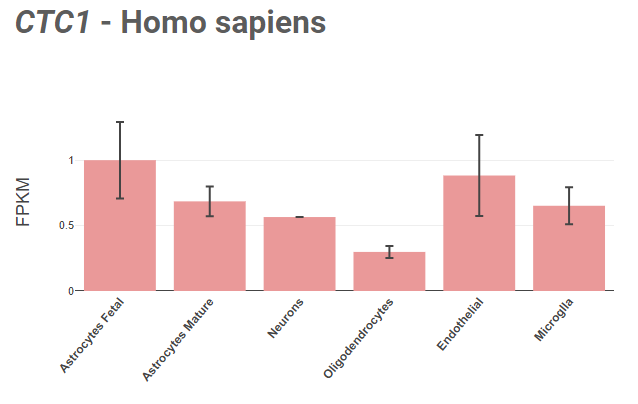

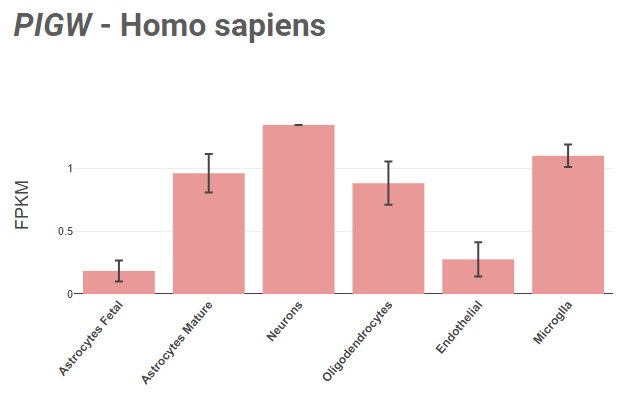

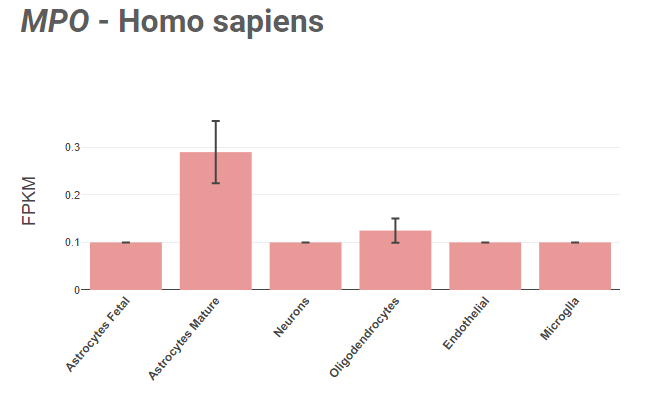

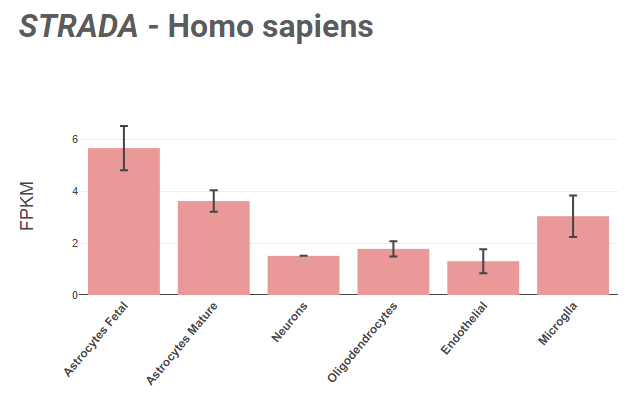

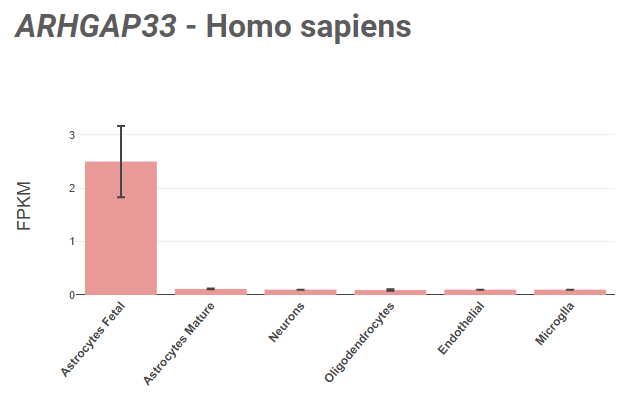

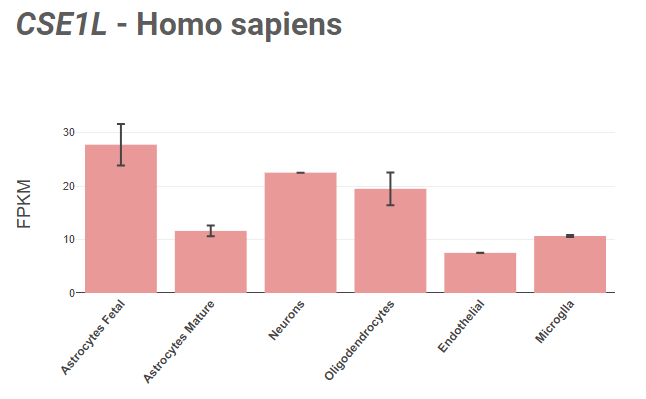

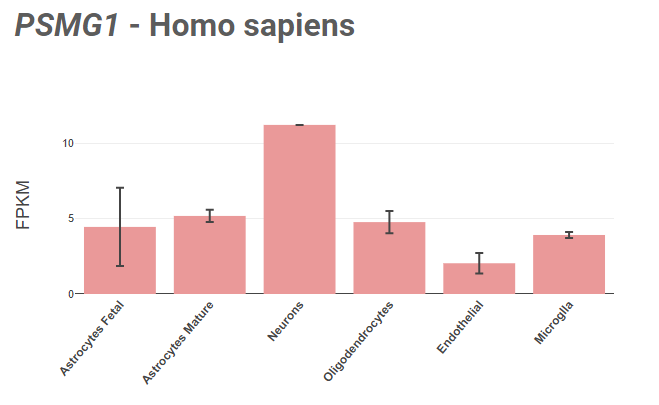

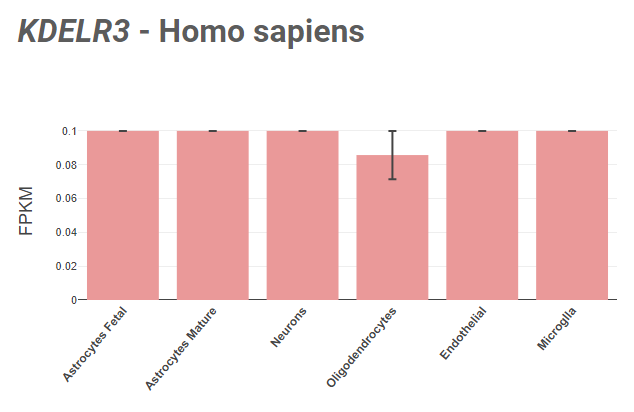

**Supplementary Figure 8**

Single-cell RNA sequencing investigation (Brain RNA-Seq) of the genes mapped from the shared loci between childhood absence epilepsy and general cognitive ability shows that the mapped genes are significantly expressed in brain cells.

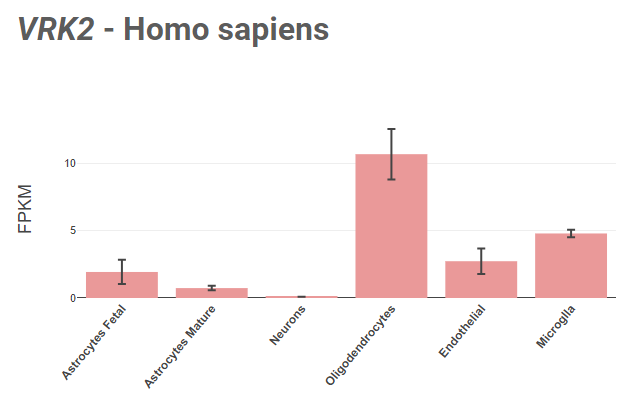

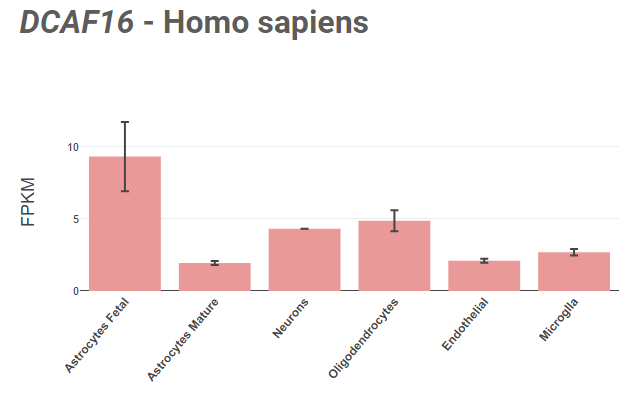

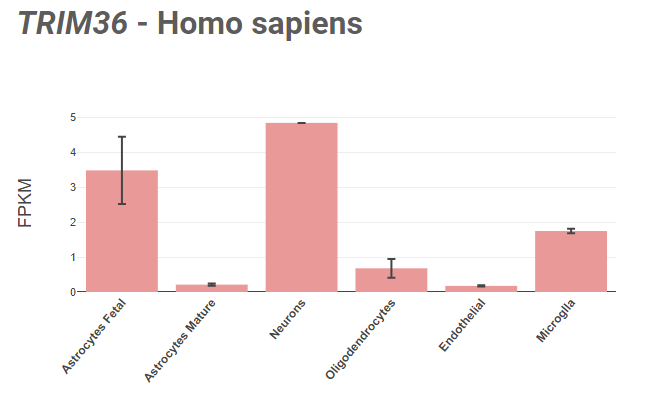

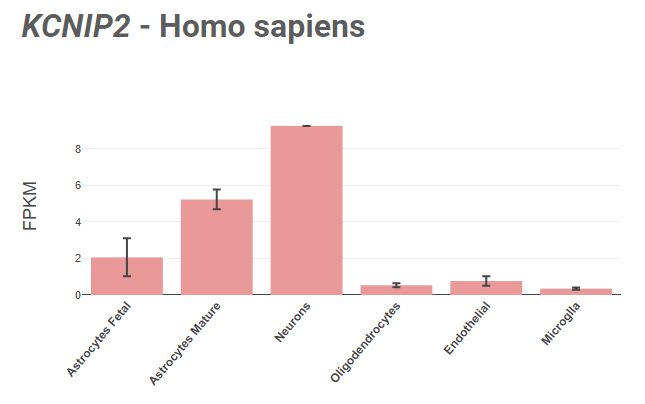

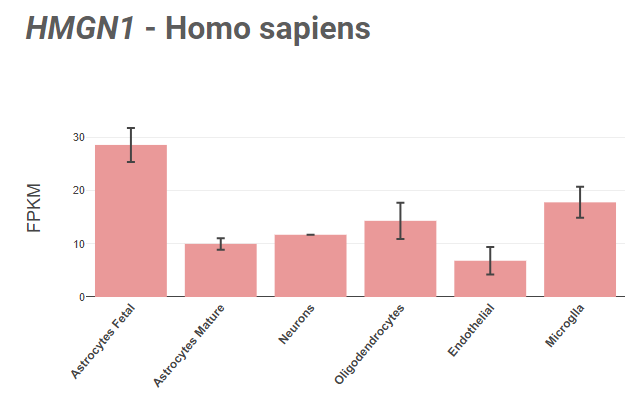

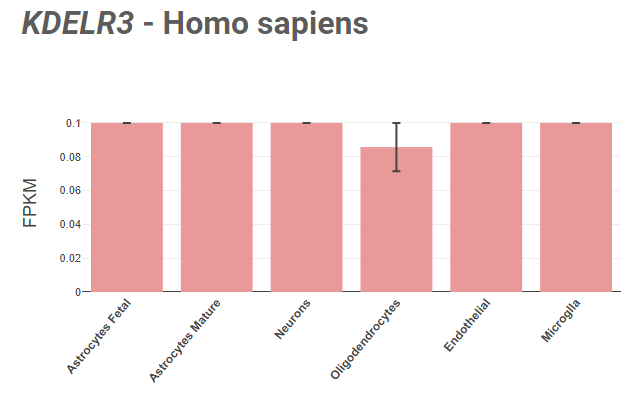

**Supplementary Figure 9**

Single-cell RNA sequencing investigation (Brain RNA-Seq) of the genes mapped from the shared loci between juvenile myoclonic epilepsy and general cognitive ability shows that the mapped genes are significantly expressed in brain cells.

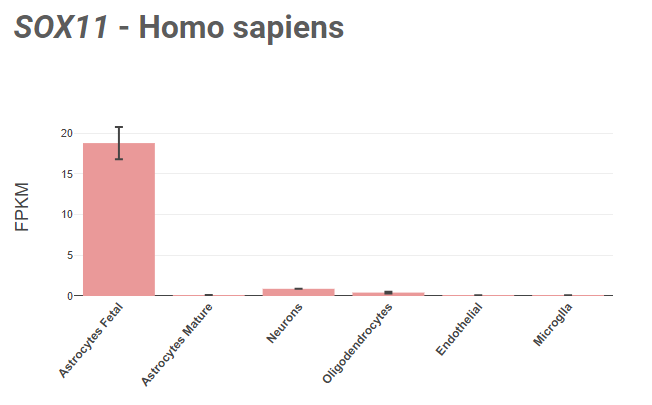

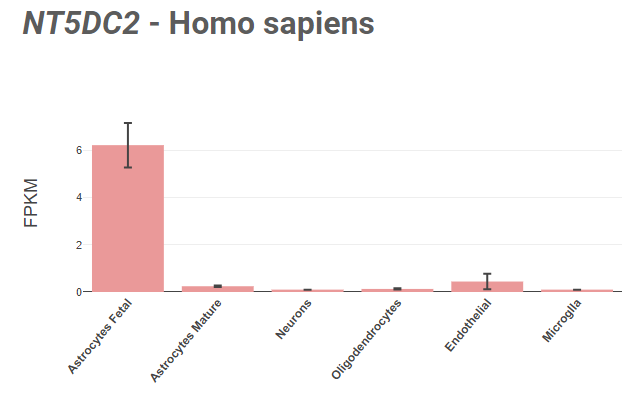

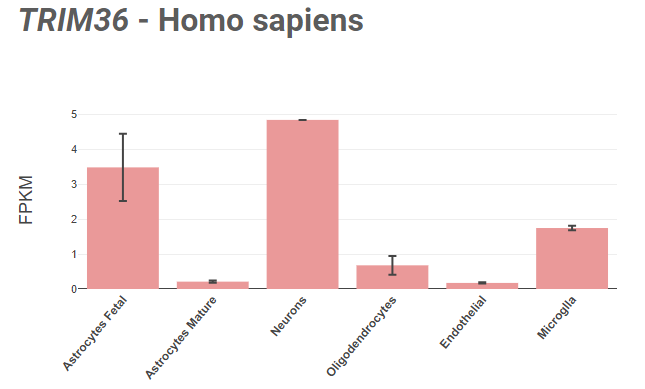

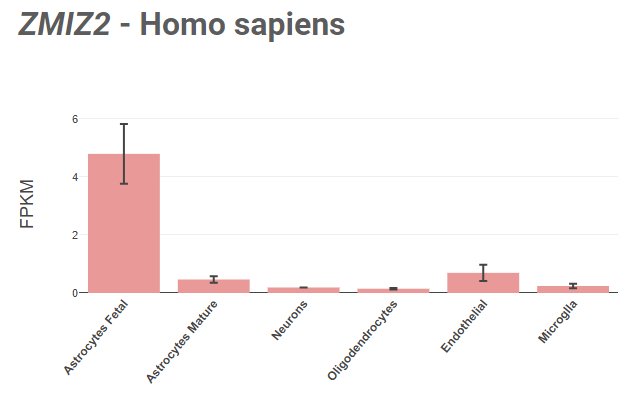

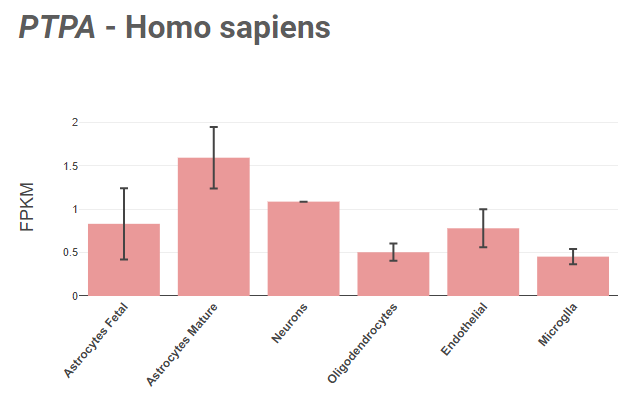

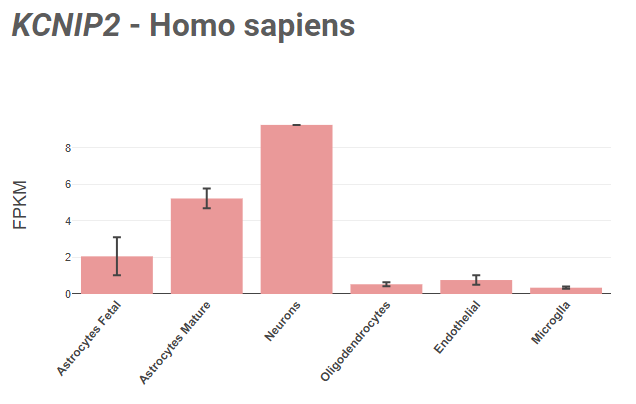

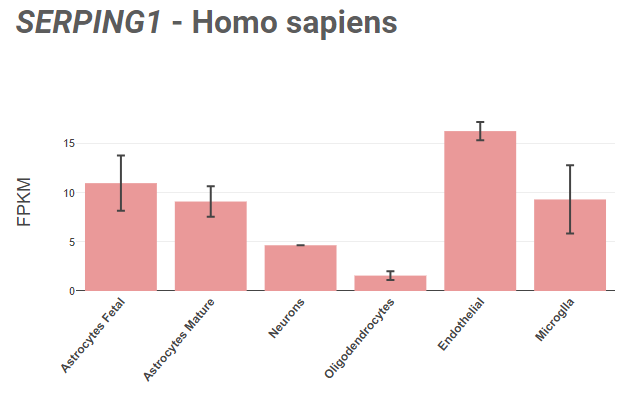

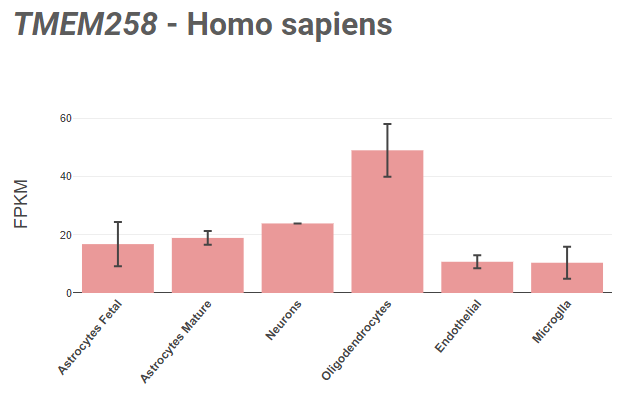

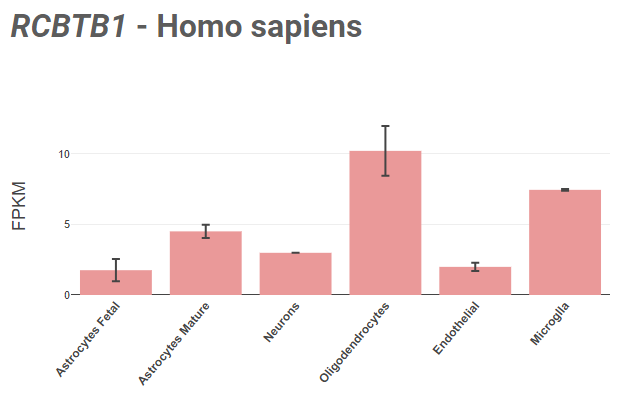

**Supplementary Table A**

**Overview of the mapped genes from the novel epilepsy loci.**

| **Gene Name** | **Summary by NCBI^2^** |
| --- | --- |
| **Shared between GGE & COG** | |
| TCEA3 | transcription elongation factor A3  Official Symbol: TCEA3  Other Aliases: TFIIS, TFIIS.H  Location: 1p36.12  Summary  Predicted to enable DNA binding activity and zinc ion binding activity. Predicted to be involved in regulation of transcription, DNA-templated and transcription, DNA-templated. Predicted to be located in nucleus. [provided by Alliance of Genome Resources, Apr 2022] |
| CACNA1E | calcium voltage-gated channel subunit alpha1 E  Official Symbol: CACNA1E  Other Aliases: BII, CACH6, CACNL1A6, Cav2.3, DEE69, EIEE69, gm139  Location: 1q25.3  Summary  Voltage-dependent calcium channels are multisubunit complexes consisting of alpha-1, alpha-2, beta, and delta subunits in a 1:1:1:1 ratio. These channels mediate the entry of calcium ions into excitable cells, and are also involved in a variety of calcium-dependent processes, including muscle contraction, hormone or neurotransmitter release, gene expression, cell motility, cell division and cell death. This gene encodes the alpha-1E subunit of the R-type calcium channels, which belong to the 'high-voltage activated' group that may be involved in the modulation of firing patterns of neurons important for information processing. Alternatively spliced transcript variants encoding different isoforms have been described for this gene. [provided by RefSeq, Apr 2011] |
| BCL11A | BCL11 transcription factor A  Official Symbol: BCL11A  Other Aliases: CTIP1, DILOS, EVI9, HBFQTL5, SMARCM1, ZNF856  Location: 2p16.1  Summary This gene encodes a C2H2 type zinc-finger protein by its similarity to the mouse Bcl11a/Evi9 protein. The corresponding mouse gene is a common site of retroviral integration in myeloid leukemia, and may function as a leukemia disease gene, in part, through its interaction with BCL6. During hematopoietic cell differentiation, this gene is down-regulated. It is possibly involved in lymphoma pathogenesis since translocations associated with B-cell malignancies also deregulates its expression. Multiple transcript variants encoding several different isoforms have been found for this gene. [provided by RefSeq, Jul 2008] |
| MITF | melanocyte inducing transcription factor  Official Symbol: MITF  Other Aliases: CMM8, COMMAD, MI-A, WS2, WS2A, bHLHe32, MITF  Location: 3p13  Summary The protein encoded by this gene is a transcription factor that contains both basic helix-loop-helix and leucine zipper structural features. The encoded protein regulates melanocyte development and is responsible for pigment cell-specific transcription of the melanogenesis enzyme genes. Heterozygous mutations in the this gene cause auditory-pigmentary syndromes, such as Waardenburg syndrome type 2 and Tietz syndrome. [provided by RefSeq, Aug 2017] |
| PPA2 | inorganic pyrophosphatase 2  Official Symbol: PPA2  Other Aliases: HSPC124, SCFAI, SCFI, SID6-306  Location: 4q24  Summary The protein encoded by this gene is localized to the mitochondrion, is highly similar to members of the inorganic pyrophosphatase (PPase) family and contains the signature sequence essential for the catalytic activity of PPase. PPases catalyze the hydrolysis of pyrophosphate to inorganic phosphate, which is important for the phosphate metabolism of cells. Alternate transcriptional splice variants, encoding different isoforms, have been characterized. [provided by RefSeq, Jul 2008] |
| GATB | glutamyl-tRNA amidotransferase subunit B  Official Symbol: GATB  Other Aliases: COXPD41, HSPC199, PET112, PET112L  Location: 4q31.3  Summary Enables glutaminyl-tRNA synthase (glutamine-hydrolyzing) activity. Involved in glutaminyl-tRNAGln biosynthesis via transamidation and mitochondrial translation. Located in mitochondrion. Part of glutamyl-tRNA(Gln) amidotransferase complex. Implicated in combined oxidative phosphorylation deficiency 41. [provided by Alliance of Genome Resources, Apr 2022] |
| GAPT | GRB2 binding adaptor protein, transmembrane  Official Symbol: GAPT  Other Aliases: C5orf29  Location: 5q11.2  Summary Predicted to be involved in B cell homeostasis and B cell proliferation involved in immune response. Located in Golgi apparatus and plasma membrane. [provided by Alliance of Genome Resources, Apr 2022] |
| SP4 | Sp4 transcription factor  Official Symbol: SP4  Other Aliases: HF1B, SPR-1  Location: 7p15.3  Summary The protein encoded by this gene is a transcription factor that can bind to the GC promoter region of a variety of genes, including those of the photoreceptor signal transduction system. The encoded protein binds to the same sites in promoter CpG islands as does the transcription factor SP1, although its expression is much more restricted compared to that of SP1. This gene may be involved in bipolar disorder and schizophrenia. [provided by RefSeq, May 2016] |
| ZMIZ2 | zinc finger MIZ-type containing 2  Official Symbol: ZMIZ2  Other Aliases: NET27, TRAFIP20, ZIMP7, hZIMP7  Location: 7p13  Summary ZMIZ2 and ZMIZ1 (MIM 607159) are members of a PIAS (see MIM 603566)-like family of proteins that interact with nuclear hormone receptors. ZMIZ2 interacts with androgen receptor (AR; MIM 313700) and enhances AR-mediated transcription (Huang et al., 2005 [PubMed 16051670]). [supplied by OMIM, May 2010] |
| MAGI2 | membrane associated guanylate kinase, WW and PDZ domain containing 2  Official Symbol: MAGI2  Other Aliases: ACVRIP1, AIP-1, AIP1, ARIP1, MAGI-2, NPHS15, SSCAM  Location: 7q21.11  Summary The protein encoded by this gene interacts with atrophin-1. Atrophin-1 contains a polyglutamine repeat, expansion of which is responsible for dentatorubral and pallidoluysian atrophy. This encoded protein is characterized by two WW domains, a guanylate kinase-like domain, and multiple PDZ domains. It has structural similarity to the membrane-associated guanylate kinase homologue (MAGUK) family. [provided by RefSeq, Jul 2008] |
| SLC24A2 | solute carrier family 24 member 2  Official Symbol: SLC24A2  Other Aliases: NCKX2  Location: 9p22.1-p21.3  Summary  This gene encodes a member of the calcium/cation antiporter superfamily of transport proteins. The encoded protein belongs to the SLC24 branch of exchangers, which can mediate the extrusion of one Ca2+ ion and one K+ ion in exchange for four Na+ ions. This family member is a retinal cone/brain exchanger that can mediate a light-induced decrease in free Ca2+ concentration. This protein may also play a neuroprotective role during ischemic brain injury. Alternative splicing results in multiple transcript variants. [provided by RefSeq, Aug 2011] |
| ELAVL2 | ELAV like RNA binding protein 2  Official Symbol: ELAVL2  Other Aliases: HEL-N1, HELN1, HUB  Location: 9p21.3  Summary  In humans, the ELAV-like RNA binding protein gene family has four members (ELAVL1-4). ELAVL RNA binding proteins recognize AU-rich elements in the 3' UTRs of gene transcripts and thereby regulate gene expression post-transcriptionally. The protein encoded by this gene binds to several 3' UTRs, including its own and also that of FOS, ID, and POU5F1. This gene encodes ELAVL2 and, like ELAVL3 and ELAVL4, is expressed specifically in neurons and primarily localizes to the cytoplasm. This protein also forms a cytosolic complex with the normally nuclear-localized ELAVL1 protein. Alternative splicing of this gene results in multiple transcript variants encoding distinct protein isoforms. [provided by RefSeq, Jul 2020] |
| CCDC6 | coiled-coil domain containing 6  Official Symbol: CCDC6  Other Aliases: D10S170, H4, PTC, PTC1, TPC, TST1  Location: 10q21.2  Summary  This gene encodes a coiled-coil domain-containing protein. The encoded protein is ubiquitously expressed and may function as a tumor suppressor. A chromosomal rearrangement resulting in the expression of a fusion gene containing a portion of this gene and the intracellular kinase-encoding domain of the ret proto-oncogene is the cause of thyroid papillary carcinoma.[provided by RefSeq, Sep 2010] |
| FBXO3 | F-box protein 3  Official Symbol: FBXO3  Other Aliases: FBA, FBX3  Location: 11p13  Summary  This gene encodes a member of the F-box protein family which is characterized by an approximately 40 amino acid motif, the F-box. The F-box proteins constitute one of the four subunits of the ubiquitin protein ligase complex called SCFs (SKP1-cullin-F-box), which function in phosphorylation-dependent ubiquitination. The F-box proteins are divided into 3 classes: Fbws containing WD-40 domains, Fbls containing leucine-rich repeats, and Fbxs containing either different protein-protein interaction modules or no recognizable motifs. The protein encoded by this gene belongs to the Fbxs class. Alternative splicing of this gene generates 2 transcript variants diverging at the 3' end. [provided by RefSeq, Jul 2008] |
| NCAM1 | neural cell adhesion molecule 1  Official Symbol: NCAM1  Other Aliases: CD56, MSK39, NCAM  Location: 11q23.2  Summary  This gene encodes a cell adhesion protein which is a member of the immunoglobulin superfamily. The encoded protein is involved in cell-to-cell interactions as well as cell-matrix interactions during development and differentiation. The encoded protein plays a role in the development of the nervous system by regulating neurogenesis, neurite outgrowth, and cell migration. This protein is also involved in the expansion of T lymphocytes, B lymphocytes and natural killer (NK) cells which play an important role in immune surveillance. This protein plays a role in signal transduction by interacting with fibroblast growth factor receptors, N-cadherin and other components of the extracellular matrix and by triggering signalling cascades involving FYN-focal adhesion kinase (FAK), mitogen-activated protein kinase (MAPK), and phosphatidylinositol 3-kinase (PI3K). One prominent isoform of this gene, cell surface molecule CD56, plays a role in several myeloproliferative disorders such as acute myeloid leukemia and differential expression of this gene is associated with differential disease progression. For example, increased expression of CD56 is correlated with lower survival in acute myeloid leukemia patients whereas increased severity of COVID-19 is correlated with decreased abundance of CD56-expressing NK cells in peripheral blood. Alternative splicing results in multiple transcript variants encoding distinct protein isoforms. [provided by RefSeq, Aug 2020] |
| RAB5B | RAB5B, member RAS oncogene family  Official Symbol: RAB5B  Location: 12q13.2  Summary  Enables GDP binding activity; GTP-dependent protein binding activity; and GTPase activity. Involved in antigen processing and presentation and plasma membrane to endosome transport. Located in endosome and extracellular exosome. [provided by Alliance of Genome Resources, Apr 2022] |
| CABP1 | calcium binding protein 1  Official Symbol: CABP1  Other Aliases: CALBRAIN, HCALB_BR  Location: 12q24.31  Summary  Calcium binding proteins are an important component of calcium mediated cellular signal transduction. This gene encodes a protein that belongs to a subfamily of calcium binding proteins which share similarity to calmodulin. The protein encoded by this gene regulates the gating of voltage-gated calcium ion channels. This protein inhibits calcium-dependent inactivation and supports calcium-dependent facilitation of ion channels containing voltage-dependent L-type calcium channel subunit alpha-1C. This protein also regulates calcium-dependent activity of inositol 1,4,5-triphosphate receptors, P/Q-type voltage-gated calcium channels, and transient receptor potential channel TRPC5. This gene is predominantly expressed in retina and brain. Alternative splicing results in multiple transcript variants encoding disinct isoforms. [provided by RefSeq, Jul 2012] |
| NOVA1 | NOVA alternative splicing regulator 1  Official Symbol: NOVA1  Other Aliases: Nova-1  Location: 14q12  Summary  This gene encodes a neuron-specific RNA-binding protein, a member of the Nova family of paraneoplastic disease antigens, that is recognized and inhibited by paraneoplastic antibodies. These antibodies are found in the sera of patients with paraneoplastic opsoclonus-ataxia, breast cancer, and small cell lung cancer. Alternatively spliced transcripts encoding distinct isoforms have been described. [provided by RefSeq, Jul 2008] |
| ERCC4 | ERCC excision repair 4, endonuclease catalytic subunit  Official Symbol: ERCC4  Other Aliases: ERCC11, FANCQ, RAD1, XFEPS, XPF  Location: 16p13.12  Summary  The protein encoded by this gene forms a complex with ERCC1 and is involved in the 5' incision made during nucleotide excision repair. This complex is a structure specific DNA repair endonuclease that interacts with EME1. Defects in this gene are a cause of xeroderma pigmentosum complementation group F (XP-F), or xeroderma pigmentosum VI (XP6).[provided by RefSeq, Mar 2009] |
| SLC5A11 | solute carrier family 5 member 11  Official Symbol: SLC5A11  Other Aliases: KST1, RKST1, SGLT6, SMIT2  Location: 16p12.1  Summary  Cotransporters, such as SLC5A11, represent a major class of proteins that make use of ion gradients to drive active transport for the cellular accumulation of nutrients, neurotransmitters, osmolytes, and ions Roll et al. (2002) [PubMed 12039040]. [supplied by OMIM, Mar 2008] |
| MPO | myeloperoxidase  Official Symbol: MPO  Location: 17q22  Summary  Myeloperoxidase (MPO) is a heme protein synthesized during myeloid differentiation that constitutes the major component of neutrophil azurophilic granules. Produced as a single chain precursor, myeloperoxidase is subsequently cleaved into a light and heavy chain. The mature myeloperoxidase is a tetramer composed of 2 light chains and 2 heavy chains. This enzyme produces hypohalous acids central to the microbicidal activity of neutrophils. [provided by RefSeq, Nov 2014] |
| STRADA | STE20 related adaptor alpha  Official Symbol: STRADA  Other Aliases: LYK5, NY-BR-96, PMSE, STRAD, STRAD alpha, Stlk  Location: 17q23.3  Summary  The protein encoded by this gene contains a STE20-like kinase domain, but lacks several residues that are critical for catalytic activity, so it is termed a 'pseudokinase'. The protein forms a heterotrimeric complex with serine/threonine kinase 11 (STK11, also known as LKB1) and the scaffolding protein calcium binding protein 39 (CAB39, also known as MO25). The protein activates STK11 leading to the phosphorylation of both proteins and excluding STK11 from the nucleus. The protein is necessary for STK11-induced G1 cell cycle arrest. A mutation in this gene has been shown to result in polyhydramnios, megalencephaly, and symptomatic epilepsy (PMSE) syndrome. Multiple transcript variants encoding different isoforms have been found for this gene. Additional transcript variants have been described but their full-length nature is not known. [provided by RefSeq, Sep 2009] |
| CSE1L | chromosome segregation 1 like  Official Symbol: CSE1L  Other Aliases: CAS, CSE1, XPO2  Location: 20q13.13  Summary  Proteins that carry a nuclear localization signal (NLS) are transported into the nucleus by the importin-alpha/beta heterodimer. Importin-alpha binds the NLS, while importin-beta mediates translocation through the nuclear pore complex. After translocation, RanGTP binds importin-beta and displaces importin-alpha. Importin-alpha must then be returned to the cytoplasm, leaving the NLS protein behind. The protein encoded by this gene binds strongly to NLS-free importin-alpha, and this binding is released in the cytoplasm by the combined action of RANBP1 and RANGAP1. In addition, the encoded protein may play a role both in apoptosis and in cell proliferation. Alternatively spliced transcript variants have been found for this gene. [provided by RefSeq, Jan 2012] |
| PSMG1 | proteasome assembly chaperone 1  Official Symbol: PSMG1  Other Aliases: C21LRP, DSCR2, LRPC21, PAC-1, PAC1  Location: 21q22.2  Summary  Enables molecular adaptor activity. Involved in chaperone-mediated protein complex assembly. Located in several cellular components, including Golgi apparatus; endoplasmic reticulum; and nucleoplasm. Part of chaperone complex. [provided by Alliance of Genome Resources, Apr 2022] |
| KDELR3 | KDEL endoplasmic reticulum protein retention receptor 3  Official Symbol: KDELR3  Other Aliases: ERD23, ERD2L3  Location: 22q13.1  Summary  This gene encodes a member of the KDEL endoplasmic reticulum protein retention receptor family. Retention of resident soluble proteins in the lumen of the endoplasmic reticulum (ER) is achieved in both yeast and animal cells by their continual retrieval from the cis-Golgi, or a pre-Golgi compartment. Sorting of these proteins is dependent on a C-terminal tetrapeptide signal, usually lys-asp-glu-leu (KDEL) in animal cells, and his-asp-glu-leu (HDEL) in S. cerevisiae. This process is mediated by a receptor that recognizes, and binds the tetrapeptide-containing protein, and returns it to the ER. In yeast, the sorting receptor encoded by a single gene, ERD2, is a seven-transmembrane protein. Unlike yeast, several human homologs of the ERD2 gene, constituting the KDEL receptor gene family, have been described. KDELR3 was the third member of the family to be identified. Alternate splicing results in multiple transcript variants. [provided by RefSeq, Jul 2013] |
| MRTFA | myocardin related transcription factor A  Official Symbol: MRTFA  Other Aliases: BSAC, MAL, MKL, MKL1, MRTF-A  Location: 22q13.1-q13.2  Summary  The protein encoded by this gene interacts with the transcription factor myocardin, a key regulator of smooth muscle cell differentiation. The encoded protein is predominantly nuclear and may help transduce signals from the cytoskeleton to the nucleus. This gene is involved in a specific translocation event that creates a fusion of this gene and the RNA-binding motif protein-15 gene. This translocation has been associated with acute megakaryocytic leukemia. Alternative splicing results in multiple transcript variants. [provided by RefSeq, Sep 2013] |
| **Shared between ‘All epilepsy’ & COG** | |
| CR1 | complement C3b/C4b receptor 1 (Knops blood group)  Official Symbol: CR1  Other Aliases: C3BR, C4BR, CD35, KN  Location: 1q32.2  Summary  This gene is a member of the receptors of complement activation (RCA) family and is located in the 'cluster RCA' region of chromosome 1. The genome is polymorphic at this locus with allele-specific splice variants encoding different isoforms, based on the presence/absence of long homologous repeats (LHRs). The gene encodes a monomeric single-pass type I membrane glycoprotein found on erythrocytes, leukocytes, glomerular podocytes, and splenic follicular dendritic cells. The Knops blood group system is a system of antigens located on this protein. The protein mediates cellular binding to particles and immune complexes that have activated complement. Decreases in expression of this protein and/or mutations in this gene have been associated with gallbladder carcinomas, mesangiocapillary glomerulonephritis, systemic lupus erythematosus, sarcoidosis and Alzheimer's disease. Mutations in this gene have also been associated with a reduction in Plasmodium falciparum rosetting, conferring protection against severe malaria. [provided by RefSeq, May 2020] |
| LONRF2 | LON peptidase N-terminal domain and ring finger 2  Official Symbol: LONRF2  Other Aliases: RNF192  Location: 2q11.2  Summary  Predicted to enable metal ion binding activity. [provided by Alliance of Genome Resources, Apr 2022] |
| SLC20A1 | solute carrier family 20 member 1  Official Symbol: SLC20A1  Other Aliases: GLVR1, Glvr-1, PIT1, PiT-1  Location: 2q14.1  Summary  The protein encoded by this gene is a sodium-phosphate symporter that absorbs phosphate from interstitial fluid for use in cellular functions such as metabolism, signal transduction, and nucleic acid and lipid synthesis. The encoded protein is also a retroviral receptor, causing human cells to be susceptible to infection by gibbon ape leukemia virus, simian sarcoma-associated virus, feline leukemia virus subgroup B, and 10A1 murine leukemia virus.[provided by RefSeq, Mar 2011] |
| SLC39A8 | solute carrier family 39 member 8  Official Symbol: SLC39A8  Other Aliases: BIGM103, CDG2N, LZT-Hs6, PP3105, ZIP8  Location: 4q24  Summary  This gene encodes a member of the SLC39 family of solute-carrier genes, which show structural characteristics of zinc transporters. The encoded protein is glycosylated and found in the plasma membrane and mitochondria, and functions in the cellular import of zinc at the onset of inflammation. It is also thought to be the primary transporter of the toxic cation cadmium, which is found in cigarette smoke. Multiple transcript variants encoding different isoforms have been found for this gene. Additional alternatively spliced transcript variants of this gene have been described, but their full-length nature is not known. [provided by RefSeq, Oct 2008] |
| ZSWIM6 | zinc finger SWIM-type containing 6  Official Symbol: ZSWIM6  Other Aliases: AFND, NEDMAGA  Location: 5q12.1  Summary  The protein encoded by this gene contains a zinc finger SWI2/SNF2 and MuDR (SWIM) domain. Proteins with SWIM domains have been found in a diverse number of species and are predicted to interact with DNA or proteins. Mutations in this gene result in acromelic frontonasal dysostosis. [provided by RefSeq, Apr 2017] |
| **Shared between CAE & COG** | |
| DCAF16 | DDB1 and CUL4 associated factor 16  Official Symbol: DCAF16  Other Aliases: C4orf30  Location: 4p15.31  Summary  Predicted to be involved in protein ubiquitination. Part of Cul4-RING E3 ubiquitin ligase complex. [provided by Alliance of Genome Resources, Apr 2022] |
| KIFBP | kinesin family binding protein  Official Symbol: KIFBP  Other Aliases: KBP, KIAA1279, KIF1BP, TTC20  Location: 10q22.1  Summary  This gene encodes a kinesin family member 1 binding protein that is characterized by two tetratrico peptide repeats. The encoded protein localizes to the mitochondria and may be involved in regulating transport of the mitochondria. Mutations in this gene are associated with Goldberg-Shprintzen megacolon syndrome. [provided by RefSeq, Mar 2010] |
| IQCN | IQ motif containing N  Official Symbol: IQCN  Other Aliases: KIAA1683, SPGF78  Location: 19p13.11  Summary  Located in mitochondrion. [provided by Alliance of Genome Resources, Apr 2022] |
| HMGN1 | high mobility group nucleosome binding domain 1  Official Symbol: HMGN1  Other Aliases: HMG14  Location: 21q22.2  Summary  The protein encoded by this gene binds nucleosomal DNA and is associated with transcriptionally active chromatin. Along with a similar protein, HMG17, the encoded protein may help maintain an open chromatin configuration around transcribable genes. [provided by RefSeq, Aug 2011] |
| KDELR3 | KDEL endoplasmic reticulum protein retention receptor 3  Official Symbol: KDELR3  Other Aliases: ERD23, ERD2L3  Location: 22q13.1  Summary  This gene encodes a member of the KDEL endoplasmic reticulum protein retention receptor family. Retention of resident soluble proteins in the lumen of the endoplasmic reticulum (ER) is achieved in both yeast and animal cells by their continual retrieval from the cis-Golgi, or a pre-Golgi compartment. Sorting of these proteins is dependent on a C-terminal tetrapeptide signal, usually lys-asp-glu-leu (KDEL) in animal cells, and his-asp-glu-leu (HDEL) in S. cerevisiae. This process is mediated by a receptor that recognizes, and binds the tetrapeptide-containing protein, and returns it to the ER. In yeast, the sorting receptor encoded by a single gene, ERD2, is a seven-transmembrane protein. Unlike yeast, several human homologs of the ERD2 gene, constituting the KDEL receptor gene family, have been described. KDELR3 was the third member of the family to be identified. Alternate splicing results in multiple transcript variants. [provided by RefSeq, Jul 2013] |
| **Shared between JME & COG** | |
| SOX11 | SRY-box transcription factor 11  Official Symbol: SOX11  Other Aliases: CSS9, IDDMOH, MRD27  Location: 2p25.2  Summary  This intronless gene encodes a member of the SOX (SRY-related HMG-box) family of transcription factors involved in the regulation of embryonic development and in the determination of the cell fate. The encoded protein may act as a transcriptional regulator after forming a protein complex with other proteins. The protein may function in the developing nervous system and play a role in tumorigenesis. [provided by RefSeq, Jul 2008] |
| ZMIZ2 | zinc finger MIZ-type containing 2  Official Symbol: ZMIZ2  Other Aliases: NET27, TRAFIP20, ZIMP7, hZIMP7  Location: 7p13  Summary ZMIZ2 and ZMIZ1 (MIM 607159) are members of a PIAS (see MIM 603566)-like family of proteins that interact with nuclear hormone receptors. ZMIZ2 interacts with androgen receptor (AR; MIM 313700) and enhances AR-mediated transcription (Huang et al., 2005 [PubMed 16051670]).[supplied by OMIM, May 2010] |
| PTPA | protein phosphatase 2 phosphatase activator  Official Symbol: PTPA  Other Aliases: PARK25, PP2A, PPP2R4, PR53  Location: 9q34.11  Summary  Protein phosphatase 2A is one of the four major Ser/Thr phosphatases and is implicated in the negative control of cell growth and division. Protein phosphatase 2A holoenzymes are heterotrimeric proteins composed of a structural subunit A, a catalytic subunit C, and a regulatory subunit B. The regulatory subunit is encoded by a diverse set of genes that have been grouped into the B/PR55, B'/PR61, and B''/PR72 families. These different regulatory subunits confer distinct enzymatic specificities and intracellular localizations to the holozenzyme. The product of this gene belongs to the B' family. This gene encodes a specific phosphotyrosyl phosphatase activator of the dimeric form of protein phosphatase 2A. Alternative splicing results in multiple transcript variants encoding different isoforms. [provided by RefSeq, Jul 2008] |
| SERPING1 | serpin family G member 1  Official Symbol: SERPING1  Other Aliases: C1IN, C1INH, C1NH, HAE1, HAE2  Location: 11q12.1  Summary  This gene encodes a highly glycosylated plasma protein involved in the regulation of the complement cascade. Its encoded protein, C1 inhibitor, inhibits activated C1r and C1s of the first complement component and thus regulates complement activation. It is synthesized in the liver, and its deficiency is associated with hereditary angioneurotic oedema (HANE). Alternative splicing results in multiple transcript variants encoding the same isoform. [provided by RefSeq, May 2020] |

Gene summaries of the mapped genes from the novel epilepsy loci are derived from the National Center for Biotechnology Information (NCBI) database. Abbreviations: COG, general cognitive ability; All, ‘all epilepsy’; GGE, genetic generalized epilepsies; CAE, childhood absence epilepsy; JME, juvenile myoclonic epilepsy.
